# Green spaces, especially forest, linked to lower SARS-CoV-2 infection rates: A one-year nationwide study

**DOI:** 10.1101/2021.08.04.21261420

**Authors:** Bin Jiang, Yuwen Yang, Long Chen, Xueming Liu, Xueying Wu, Bin Chen, Chris Webster, William C. Sullivan, JingJing Wang, Yi Lu

## Abstract

This study examined the associations between green spaces and one-years’ worth of SARS-CoV- 2 infection rates across all 3,108 counties in the contiguous United States after controlling for multiple categories of confounding factors. We found green spaces at the county level have a significant negative association with infection rates. Among all types of green spaces, forest yields the most consistent and strongest negative association. Sensitivity analyses confirmed the negative association of forest across five urbanicity levels, and the strength of the association increases as disease incidence increases across five time periods. Although forest located in moderately urbanized counties yields the strongest association, the negative pattern of significant associations holds across all five urbanicity levels. A population-weighted analysis revealed that proximity to forest within a moderate walking distance (≤ 1.0–1.4 km) may provide the greatest protection against the risk of infection.

## 1. Introduction

The coronavirus disease 2019 (COVID-19) pandemic is an ongoing global crisis that has profoundly affected people’s health and wellbeing across the world (Holman et al., 2020; Walker et al., 2020; World Health Organization, 2021). A variety of studies have investigated the risk of infection with Severe Acute Respiratory Syndrome Coronavirus 2 (SARS-CoV-2), which causes COVID-19, from medical, social, economic, demographic, and architectural perspectives (Badr et al., 2020; Carteni et al., 2020; Mena et al., 2021; Muller et al., 2021). There is growing evidence that we have overlooked the salutary impact that nearby green spaces may have for lowering the risk of infection with SARS-CoV-2. The cost of this oversight is that we continue to create and manage built landscapes that increase the risk of infection to SARS-CoV-2 today, and that we fail to learn the extent to which land use patterns might alleviate the spread of this and other airborne infectious diseases.

Two recent studies suggest that green spaces may provide some protection against infection with SARS-CoV-2 (Lu, Chen, et al., 2021; Klompmaker, et al., 2021). The data employed in one of these studies was limited to the first wave of infections in the US that occurred from January to June, 2020 and in the other study from March to June, 2020.

Did the initially reported negative relationship found in these first two studies between green space and infection rates hold over the much larger second and third waves of the pandemic? What new can we learn about the characteristics of the green settings, population density, and the optimal buffer distance of green space exposure for reducing SARS-CoV-2 infection rates?

We consider these questions below. We begin with a review theory and published studies regarding the salutary effects that grow from exposure to green spaces and identify the gaps in our knowledge that exist after the publication of the two previous studies regarding green spaces and SARS-CoV-2. Next, we report findings from a new study designed to overcome the weakness for the previous studies and end by considering the implications of the findings for policies, regulations, and planning and design interventions that might alleviate current and future airborne pandemics.

### 1.1. Accumulating evidence: The health benefits of green spaces

The relationship between green spaces and public health is a rapidly growing field of research. In recent decades, several studies have reported on the multiple health benefits of green space exposure. The relationship between green space exposure and health outcomes has been measured at national (e.g., Lu, Chen, et al., 2021; Mitchell & Popham, 2008; Nowak et al., 2014), city (e.g., Donovan et al., 2011; Melis et al., 2015), and community/site (e.g., Chang et al., 2021; Kuo & Sullivan, 2001) scales. Green spaces, especially those in close proximity, can affect three main categories of health outcomes: mental (Jiang et al., 2014; Jiang et al., 2018), physical (Jiang, Larsen, et al., 2020; Lu, 2018; Mitchell & Popham, 2008), and social (Helbich et al., 2020; Holtan et al., 2014). The effects of green spaces on such health outcomes are complex and interdependent (Jiang, Zhang, et al., 2015; Sullivan & Bartlett, 2005). For example, green spaces can significantly reduce mental stress and fatigue, which positively influences immune function and promotes physical health (Kuo, 2015; Li et al., 2010; Parsons et al., 1998). Exposure to green spaces can also promote social cohesion and can improve physical activity levels and perceived safety. Green spaces, especially forest, can decrease air pollutants, provide pleasant visual and acoustic experiences, and create a comfortable microclimate. These critical ecological functions encourage outdoor physical activities, further enhancing mental and social health (Nowak et al., 2014; Ouyang et al., 2020; Power et al., 2015). Overall, it is widely recognized that green spaces, whether in urban or rural areas, have a significant positive effect on human health.

### 1.2. A critical knowledge gap: The relationship between green spaces and SARS-CoV-2 infection rates

The published research leaves us with a critical knowledge gap in our understanding: we do not know the extent to which exposure to green settings, especially forest, reduce SARS-CoV-2 infection risk. Two studies have attempted to investigate this relationship, however the findings were inconclusive. One study examined the 135 most urbanized counties in the contiguous United States, and found that a higher ratio of green spaces was significantly associated with a lower racial disparity in SARS-CoV-2 infection rates at the county level (Lu, Chen, et al., 2021). However, this study did not determine the association between the supply of green spaces and the general population’s infection rate across all counties in U.S., which have varying levels of urbanicity and population densities. Additionally, the infection data used in the study spanned a relatively short time period (from January to June, 2020), and excluded the moderate and peak periods of infection (from August to December, 2020).

The second study examined the association between county-level green vegetation measured by the normalized difference vegetation index (NDVI), and SARS-CoV-2 infection and death rates for 2,297 counties in the United States (Klompmaker et al., 2021). That study reported “greenness” was negatively associated with county-level SARS-CoV-2 infection rates. This study, however, had three important limitations. First, it did not identify the types of green spaces that were associated with lower infection rates. Second, the study period was relatively short (from March to June, 2020). Third, some important confounding factors were not controlled, including transportation infrastructure and services (Carteni et al., 2020; Tirachini & Cats, 2020), political and administration factors (Clinton et al., 2021), human mobility (Muller et al., 2021), commuting mode (Figueroa et al., 2021), and employment status (Mena et al., 2021).

The promise and weaknesses associated with these two previous studies demonstrate the necessity and urgency of conducting a more comprehensive assessment regarding the relationship between green spaces and SARS-CoV-2 infection rates. Understanding this relationship will enable planners and designers to develop appropriate environmental interventions to alleviate the COVID-19 pandemic, and will shed light on how we might employ the built environment to address future pandemics.

### 1.3. Research questions

In this study, we ask five related questions to uncover the relationship between green spaces and SARS-CoV-2 infection risk:

1. What is the overall association between the ratio of green spaces within a county and SARS-CoV-2 infection rate?
2. What is the relationship between different types of green spaces and SARS-CoV-2 infection rates?
3. What is the relationship between green spaces and SARS-CoV-2 infection rates across various levels of urbanicity and over distinct time-periods during the COVID-19?
4. What is the optimal buffer distance of green space exposure associated with reduced levels of SARS-CoV-2 infection rates?
5. What mechanisms might explain these relationships?

## 2. Methods

### 2.1. Overview

We investigated the association between the ratio of green spaces and SARS-CoV-2 infection rates in 3,108 counties in the contiguous United States from January 22 to December 31, 2020. We also examined the association between a variety of types of green space and infection rates in counties at five levels of urbanicity over five different time-periods. The effect of forest was determined for all 25 urbanicity–time-period combinations. Finally, in order to identify the optimal forest-buffer distance, we examined the relationship between forest located at different distances from human settlements and the infection rates within those settlements.

#### 2.1.1. Study area

We used counties, which are fundamental administrative boundaries of the United States, as the basic units of analysis, as infection data were most complete at the county level. Accordingly, 3,108 counties in the contiguous United States were included.

#### 2.1.2. SARS-CoV-2 infection rates

We collected the cumulative total number of positive cases of SARS-CoV-2 infection from the Centers for Disease Control and Prevention (CDC) and state- and local-level public health agencies (USAFACTS, 2021). The research period was from January 22 to December 31, 2020; we chose December 31 as the end point as the COVID-19 pandemic had reached its peak on approximately this week, despite some fluctuations (Fig. 1). Moreover, this end point fell prior to the rollout of large-scale vaccination programs and the presidential election, allowing us to avoid a more complex situation. The research period was further subdivided into five equal-length periods to investigate the temporal association between SARS-CoV-2 infection and green spaces. The five periods were from January 22 to March 30, March 31 to June 7, June 8 to August 15, August 16 to October 23, October 24 to December 31, 2020 (Fig. 1).

**Fig. 1.**
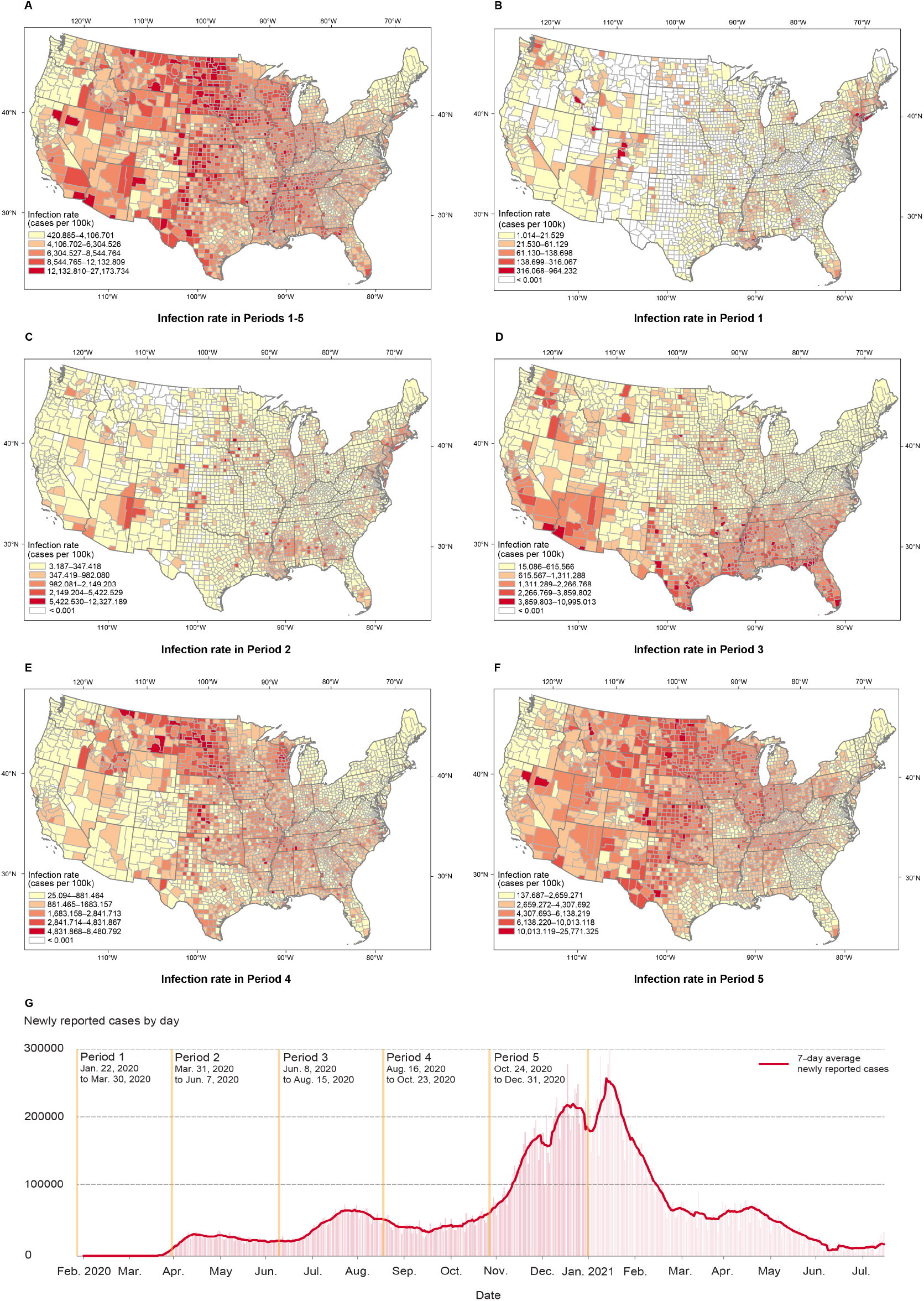
County-level severe acute respiratory syndrome coronavirus 2 (SARS-CoV-2) infection rates (cases per 100,000) in total and across the five subperiods. (A) The overall research period, from January 22 to December 31, 2020. (B) Period 1: January 22 to March 30, 2020. (C) Period 2: March 31 to June 7, 2020. (D) Period 3: June 8 to August 15, 2020. (E) Period 4: August 16 to October 23, 2020. (F) Period 5: October 24 to December 31, 2020. (G) Diagram of cases of SARS-CoV-2 from February 2020 to July 2021 in the United States (The New York Times, 2021). The COVID-19 pandemic emerged in 2020 and comprised several different periods, with low, moderate, and high infection rates. The infection rate in 2021 was significantly attenuated wide-scale vaccination, so this data was not included.

#### 2.1.3. Green spaces

We quantified four types of green space factors to reduce bias: land cover, local parks, tree canopies, and general greenness levels (Fig. 2 and Supplementary Table S1).

1. Six types of land cover with predominant natural elements were assessed at a 30-m resolution based on the National Land Cover Datasets (NLCD) (MRLC consortium, 2019): Developed open space, forest, shrub/scrub, grassland/herbaceous, hay/pasture, and cultivated crops (Fig. 3 and Table S1). The ratio of each type of land cover was calculated as the area of the land cover within the county divided by the total county land area. We also calculated population-weighted forest exposure for varying buffer distances in each county to identify an optimal distance from forest, a factor that may be critical for determining the infection rate. The population-weighted exposure incorporates population spatial distributions into green space exposure estimates by giving a proportionally greater weight to green spaces near areas of higher population density. Population-weighted forest exposure was calculated in Google Earth Engine (GEE) using the NLCD 2016. Residential population densities were estimated at a 100-m resolution for the contiguous United States using WorldPop Global Project Population Data from 2020 (Sorichetta et al., 2015). The original 30-m NLCD-based forest map was aggregated at a 100-m resolution to match that of the population data within 5 km, whereas a spatial resolution of 1 km was used for distances greater than 5 km. The population-weighted forest exposure for different buffer sizes in each county was defined by Equation 1 (Chen, Song, Jiang, et al., 2018; Chen, Song, Kwan, et al., 2018),

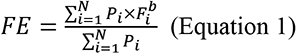

where *P_i_* represents the population of the *i*^th^ grid, 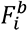 represents the forest cover of the *i*^th^ grid at a buffer size of *b* meters, *N* denotes the total number of grids for a given county, and *FE* is the estimated forest exposure level for the given county. We calculated forest exposure levels by varying the buffer size from 100 m to 50 km, to examine the association between different proximities to forest and the infection rate.
2. The data of local park were derived from the United States Parks dataset (Esri, 2021), which provides local-level details of parks and forest within the United States. The ratio of local park area was measured as the gross land area of local parks within a county divided by the total county area.
3. The percentage of tree canopy cover was estimated from the 30-m raster dataset generated by the United States Forest Service (USFS) from satellite imagery (Coulston et al., 2013; Coulston et al., 2012).
4. The overall greenness level for each county was assessed using Normalized Difference Vegetation Index (NDVI) values and GEE to integrate satellite imagery at a 30-m resolution (MRLC consortium, 2019). We used the mean NDVI value in 2020 as a proxy for overall green space exposure during the research period.

**Fig. 2.**
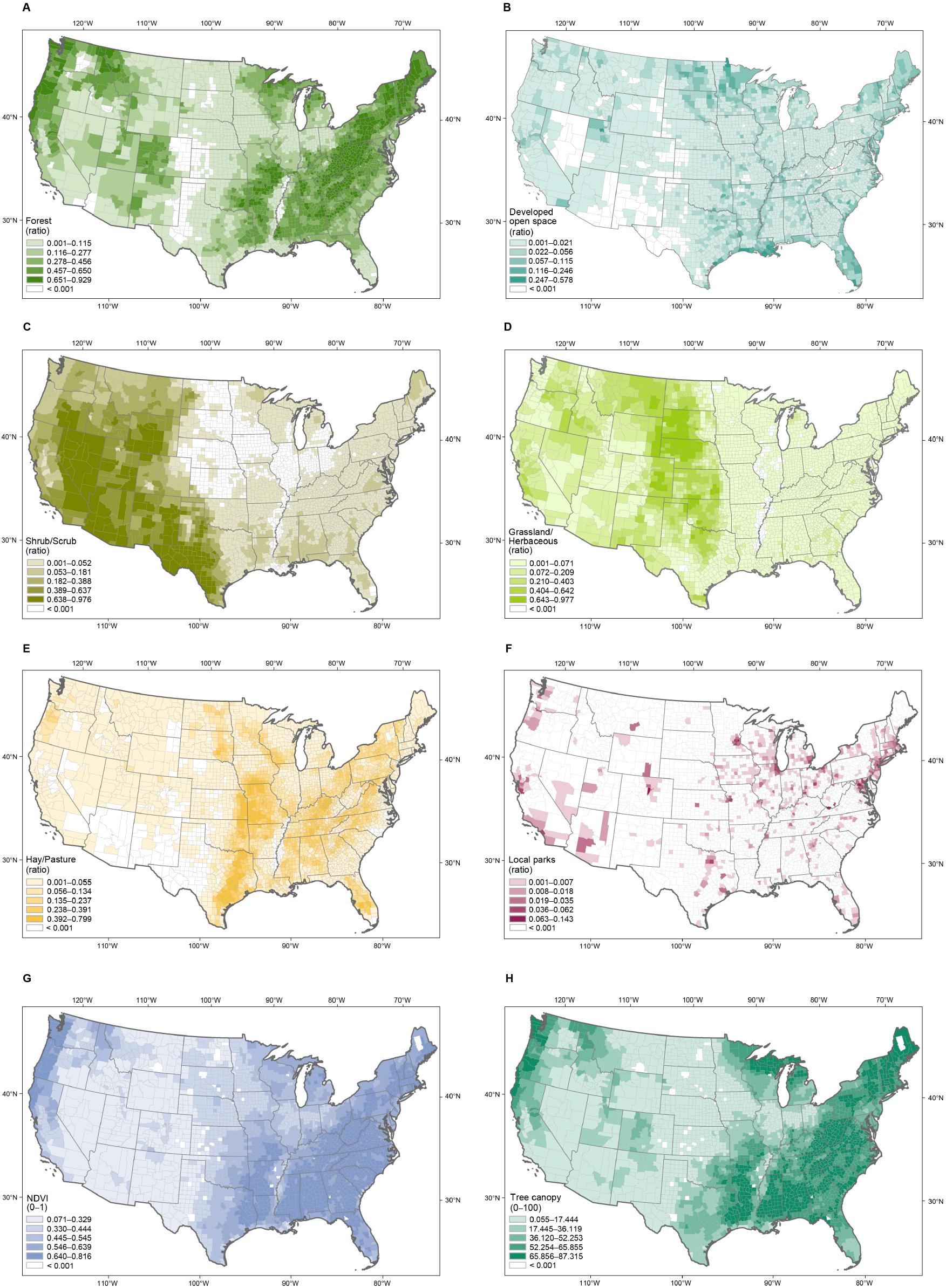
Ratio of different types of green spaces at a county level. Values represent the (A) forest, (B) developed open space, (C) shrub/scrub, (D) grassland/herbaceous, (E) hay/pasture, and (F) local parks land coverage ratios at the county level, calculated as the total relevant area divided by the entire county area. Data were extracted from the NLCD landcover dataset. Local parks land coverage was calculated based on the US Parks dataset of Esri; (G) NDVI were calculated by Google Earth Engine. (H) Tree canopy data were extracted from the NLCD landcover dataset.

**Fig. 3.**
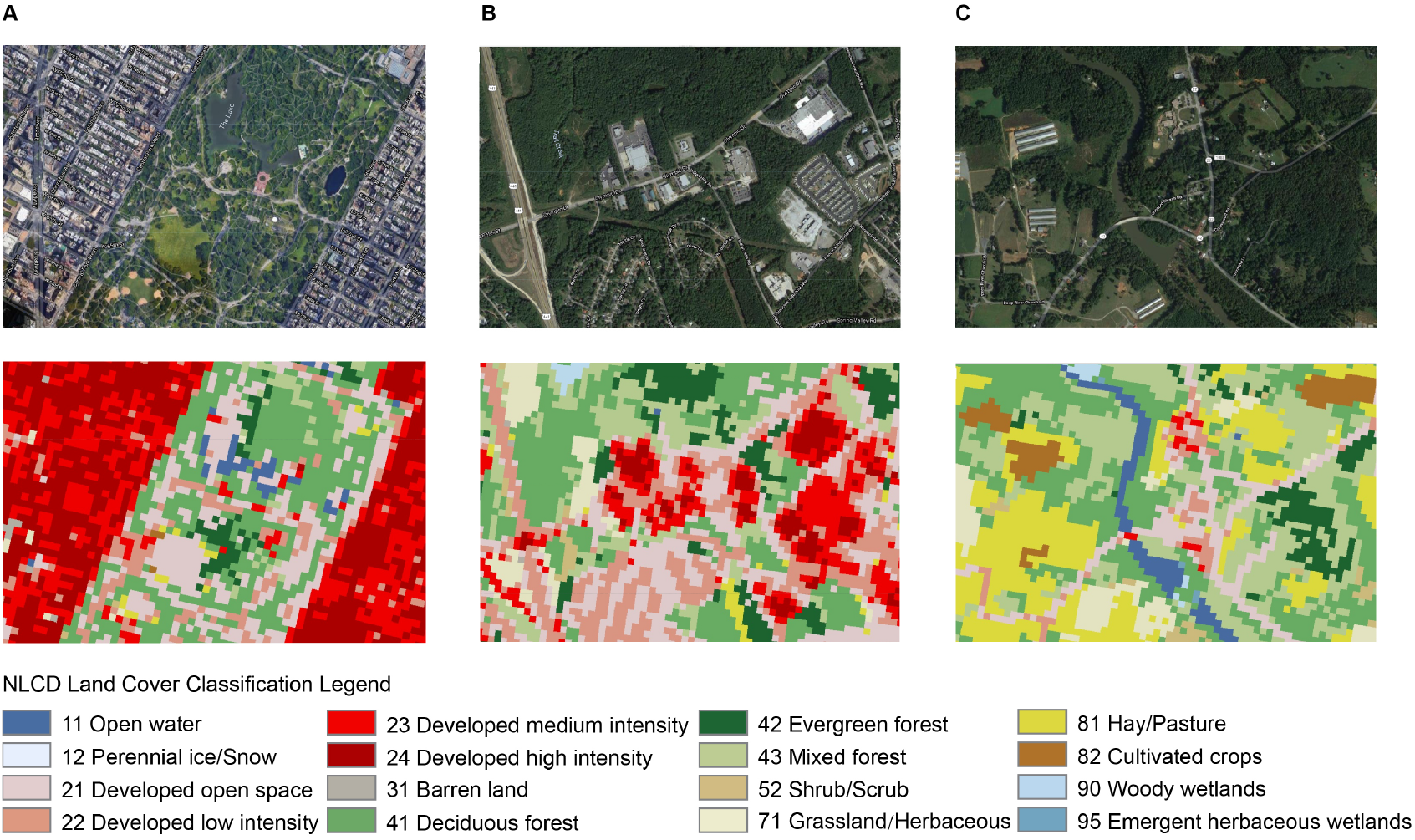
Example of land cover distributions at three different urbanicity levels. (A) New York City, NY, urbanicity level 1; (B) Athens, GA, urbanicity level 3; (C) Coleridge, NC, urbanicity level 5. Squares marked by light, moderate, and dark green colors have been identified by NLCD as forest areas, including deciduous, evergreen, and mixed forest.

#### 2.1.4. Levels of Urbanicity

Previous studies have identified an urban–rural disparity in the prevalence of SARS-CoV-2 infection in the United States (Huang et al., 2021; Pro et al., 2020). Therefore, we classified all counties into five levels of urbanicity based on the 2013 Urban-Rural Classification Scheme (NCHS, 2017), with level 1 being the most urbanized and level 5 being the most rural (Fig. S1 and Table S2).

#### 2.1.5. Confounding factors

Additionally, we adjusted for potential confounding factors in each county, including healthcare and testing rates (Wu et al., 2020), pre-existing chronic diseases (Townsend et al., 2020), and socioeconomic and demographic (Abedi et al., 2020; Clouston et al., 2021; Karaye & Horney, 2020), political (Neelon et al., 2021), behavioral (Badr et al., 2020; McGrail et al., 2020), and environmental (Chakrabarty et al., 2021; Wang et al., 2020) factors.

#### 2.1.6. Descriptive statistics of variables

The descriptive statistics for each variable entered in the regression analysis are presented in Table 1 (See Table S3 for the descriptive information for all variables).

**Table 1.** Descriptive data for SARS-CoV-2 infection rates, socioeconomic and demographic, pre-existing chronic disease, policy and regulation, behavioral, built environment, and green space factors.

| Model Predictors | Variables | Min | Max | Mean | SD | Unit or Formula |
| --- | --- | --- | --- | --- | --- | --- |
| **SARS-CoV-2 Infection rate** | Infection rate | 420.885 | 27173.734 | 6570.950 | 2762.664 | Cases per 100k |
| **Socio-economic and demographic factors** | Population density | 0.094 | 19625.842 | 98.221 | 559.972 | Persons per km<sup>2</sup> |
|  | Black non-Hispanic | 0.000 | 87.400 | 8.957 | 14.520 | Ratio |
|  | Population aged 65+ | 3.800 | 55.600 | 18.428 | 4.542 | Ratio |
|  | Gini Index | 0.257 | 0.665 | 0.446 | 0.036 | Range 0-1 |
| | Median home value | 21000.00 | 1057000.00 | 146157.40 | 89047.61 | USD\$ |
|  | Unemployment rate | 1.300 | 18.100 | 4.093 | 1.397 | Ratio |
|  | Population without high school diploma | 1.200 | 66.300 | 13.448 | 6.342 | Ratio |
| **Healthcare and testing factors** | Testing rate | 0.000 | 1711.099 | 170.197 | 107.693 | Per 100k |
|  | Population without insurance | 2.300 | 33.700 | 11.418 | 5.106 | Ratio |
| **Pre-existing chronic disease factors** | Heart failure mortality | 19.400 | 304.300 | 108.314 | 25.488 | Per 100k |
|  | Stroke mortality | 14.000 | 92.500 | 39.776 | 8.153 | Per 100k |
|  | Hypertension mortality | 19.300 | 587.300 | 131.825 | 55.366 | Per 100k |
|  | Diabetes rate | 2.200 | 28.700 | 10.506 | 3.526 | Ratio |
|  | Obesity rate | 12.30 | 57.90 | 32.76 | 5.68 | Ratio |
| **Policy and regulation factors** | State Governor Party | 0.000 | 1.000 | 0.435 | 0.496 | Democratic /Republican |
|  | Stay-at-home orders | 3.856 | 6.897 | 4.441 | 0.447 | Range 1-7 |
|  | Public mask mandates | 0.000 | 1.000 | 0.497 | 0.339 | Yes/No |
|  | Business closing and reopening | 4.781 | 8.233 | 6.331 | 0.683 | Range 1-5 |
| **Behavioral factors** | Smoker | 5.909 | 41.491 | 17.446 | 3.554 | Ratio |
|  | Essential worker | 0.178 | 0.791 | 0.526 | 0.073 | Ratio |
|  | POI visits | 0.000 | 10.507 | 1.505 | 0.904 | Per person |
|  | Commute to work by walking or bicycle | 0.000 | 42.410 | 3.427 | 3.146 | Ratio |
|  | Leisure time physical inactivity | 9.400 | 49.800 | 26.238 | 5.498 | Ratio |
|  | Mobility | 0.409 | 553.723 | 8.118 | 14.190 | Km |
|  | Mobility index | 17.156 | 947.757 | 79.520 | 40.596 | Ratio |
| **Environmental factors** | Severe housing problems | 2.700 | 39.100 | 14.308 | 4.338 | Ratio |
|  | Overcrowded Housing | 0.000 | 16.900 | 2.314 | 1.816 | Ratio |
|  | Proximity to highway | 0.000 | 16.400 | 1.943 | 1.871 | Ratio |
|  | Airport density | 0.000 | 0.039 | 0.002 | 0.002 | Number per km<sup>2</sup> |
|  | Railway density | 0.000 | 2.625 | 0.062 | 0.111 | Length of km per km<sup>2</sup> |
|  | Highway and secondary road density | 0.000 | 1.577 | 0.105 | 0.132 | Length of km per km<sup>2</sup> |
|  | PM2.5 | 1.500 | 16.000 | 7.640 | 1.674 | Kj per m<sup>2</sup> |
|  | PM10 | 7.476 | 57.922 | 17.417 | 4.755 | Ug/m<sup>3</sup> |
|  | NO<sub>2</sub> | 2.896 | 27.402 | 13.321 | 3.337 | Ppb |
|  | Average temperature | 36.809 | 79.218 | 57.687 | 7.941 | Degrees Fahrenheit |
|  | Wind speed | 3.969 | 9.923 | 7.026 | 0.800 | m/s |
| **Green space factors** | Developed open space | 0.001 | 0.337 | 0.045 | 0.039 | Developed open space area/ county area |
|  | Forest | 0.000 | 0.929 | 0.306 | 0.256 | Forest area/county area |
|  | Shrub / Scrub | 0.000 | 0.976 | 0.085 | 0.181 | Shrub and scrub area/county area |
|  | Grassland/ Herbaceous | 0.000 | 0.977 | 0.094 | 0.169 | Grassland or herbaceous area/county area |
|  | Hay / Pasture | 0.000 | 0.799 | 0.101 | 0.123 | Pasture and hay area/county area |
|  | Local parks | 0.000 | 0.143 | 0.002 | 0.009 | Local park area/county area |

### 2.2. Statistical analysis

Prior to analyses, the variance inflation factor (VIF) test was used to identify multicollinearity between the independent variables. All factors with a VIF ≥ 4 were excluded from our models (O’Brien, 2007). We employed hierarchical linear regression to address our research questions in four steps, as follows.

1. The overall associations between infection rates and green space factors were examined using two regression models. Model 1 included confounding factors only, while Model 2 included green space factors and confounding factors. This allowed us to identify the independent effect of green space factors on infection rates, while controlling for confounders.
2. We examined whether green space–infection associations varied in different time periods and at different urbanicity levels using ten regression models. Similar to the step one, for each period or urbanicity level, Model 1 included only confounding factors, while Model 2 included green space factors and confounders. Model *β*, *R*^2^, and Δ*R*^2^ values were reported.
3. The results from steps 1 and 2 revealed that forest has a consistent and strong association with infection rates. We further explored the temporality of forest–infection associations across five urbanicity levels. Regression analyses were conducted to evaluate the effect of forest on infection rates for all 25 combinations of the five time periods and five urbanicity levels. For each combination, Model 1 included all confounding factors and green space factors except forest, while Model 2 also included forest. The *β*, *R*^2^, and Δ*R*^2^ values were reported for each model (Table S4 and S5).
4. To identify the optimal distance from forest, we used a series of hierarchical linear regressions to examine the association between population-weighted forest exposure, for distances ranging from 100 m to 50 km, and the infection rate. For each buffer distance, Model 1 included all confounding factors and green space factors except forest, while Model 2 included population-weighted forest exposure. The *p* values, 95% confidence intervals (CI), and *R*^2^ and Δ*R*^2^ values were reported (Table S6).

All analyses were performed using *R*-4.0.5 with the built-in “lm” function (R Core Team, 2020).

## 3. Results

### 3.1. Overall association of green spaces with infection rates

The results of a hierarchical regression analysis are shown in Table 2. Model 1 included all confounding factors and explained 45.1% of the variability in infection rates (adjusted *R*^2^ = 0.451, *p* < 0.0001). After adding the green space factors, the overall explanatory power of Model 2 increases by 5.5% (adjusted *R^2^* = 0.506, *p* < 0.0001). The individual effects of the six types of green spaces, after controlling for confounders, revealed that forest and hay/pasture are significantly, negatively associated with overall infection rates (*p* < 0.0001) (Fig. 4). Shrub/scrub and local parks are negatively associated with infection rates at a marginally significant level (*p* < 0.05). Developed open space is significantly positively associated with infection rates (*p* <0.001). Among all types of green space, forest has the greatest effect on infection rates (*β* = −0.305).

**Fig. 4.**
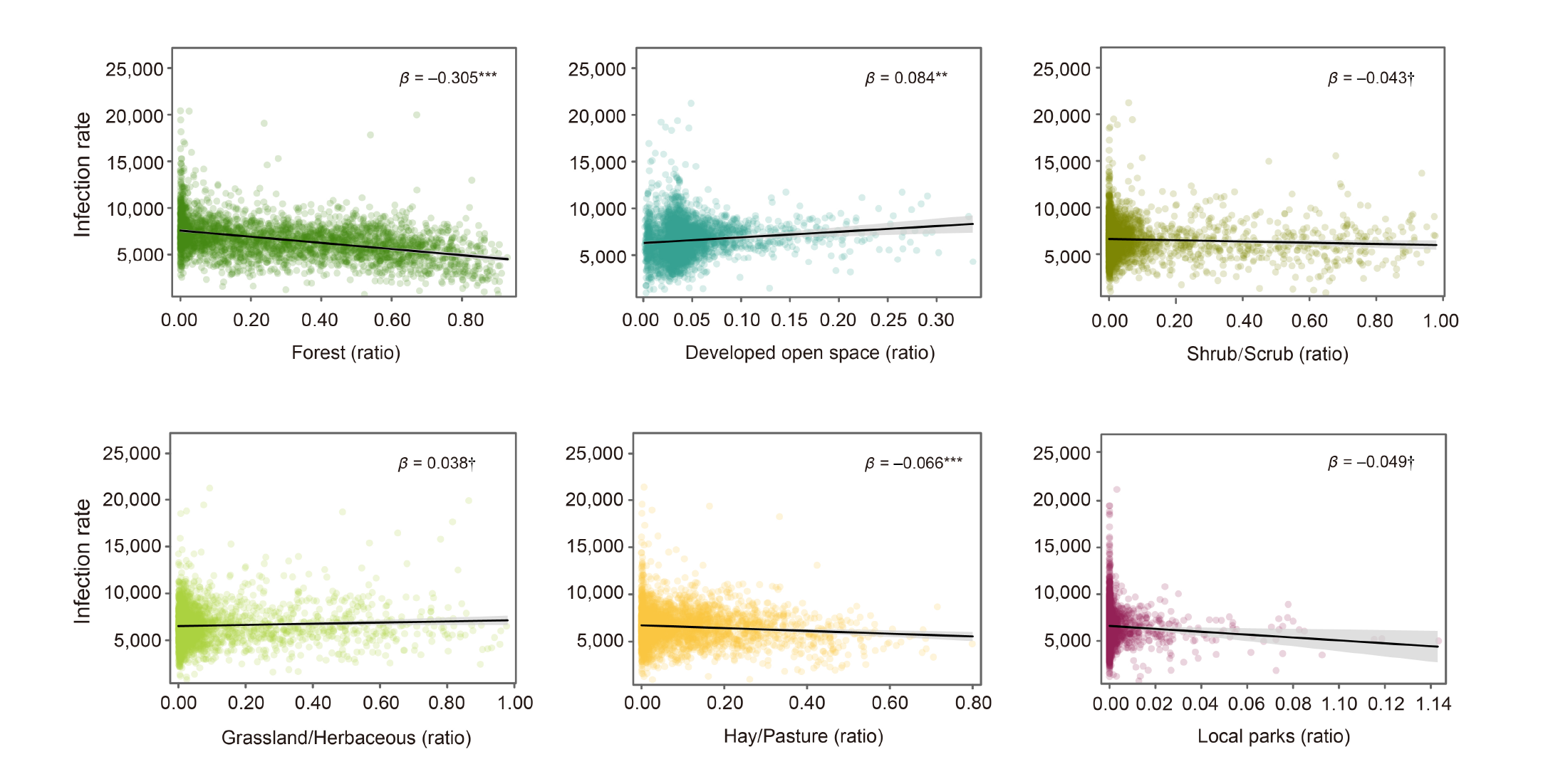
Relationships between each of the six types of green spaces and SARS-CoV-2 infection rates. Each green space factor ratio is calculated by dividing green space area by the total area of the county. Shaded areas represent pointwise 95% confidence intervals, points represent partial residuals. Lines represent the linear trend line. Note: ^ⴕ^ *p* < 0.05; * *p* < 0.01; ** *p* < 0.001; *** *p* < 0.0001.

**Table 2.**
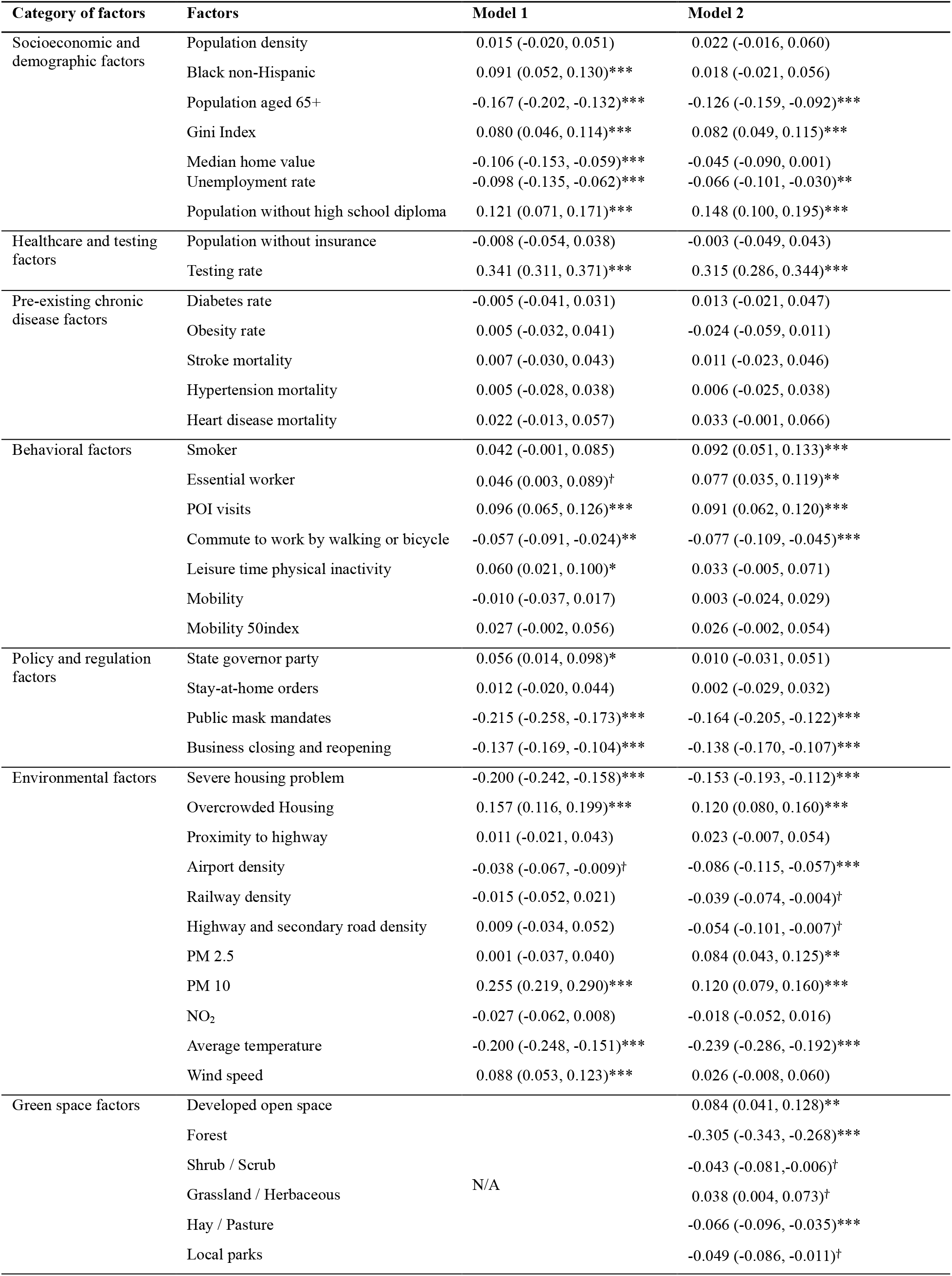
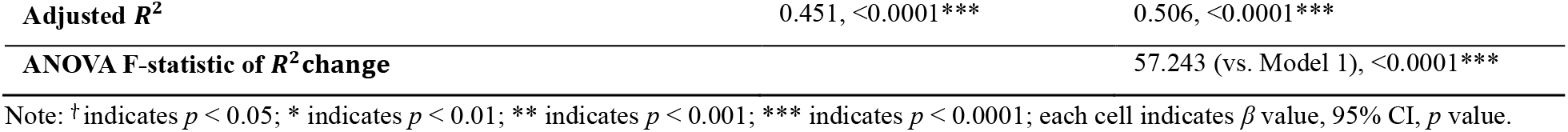
Regression analysis results of overall associations between green spaces and infection rate (N = 3108).

### 3.2. Green space-infection rate associations across levels of urbanicity

We divided all data by five levels of urbanicity, and ran separate regression models to explore the extent to which green space-infection associations varied by urbanicity (Table S4 and Fig. 5). We found that the additional explanatory power contributed by green spaces, indicated by Δ*R^2^*, is significant for all levels of urbanicity. The highest Δ*R^2^* is seen for urbanicity level 3 (9.2%), while the two extremes of urbanicity level have the lowest (level 1: 2.9%; level 5: 3.3%). As seen in Fig. 5, forest is the only type of green spaces that significantly associated with infection rates across all five levels of urbanicity, with its association being strongest at level 3 (*β* = −0.431, *p* < 0.0001).

**Fig. 5.**
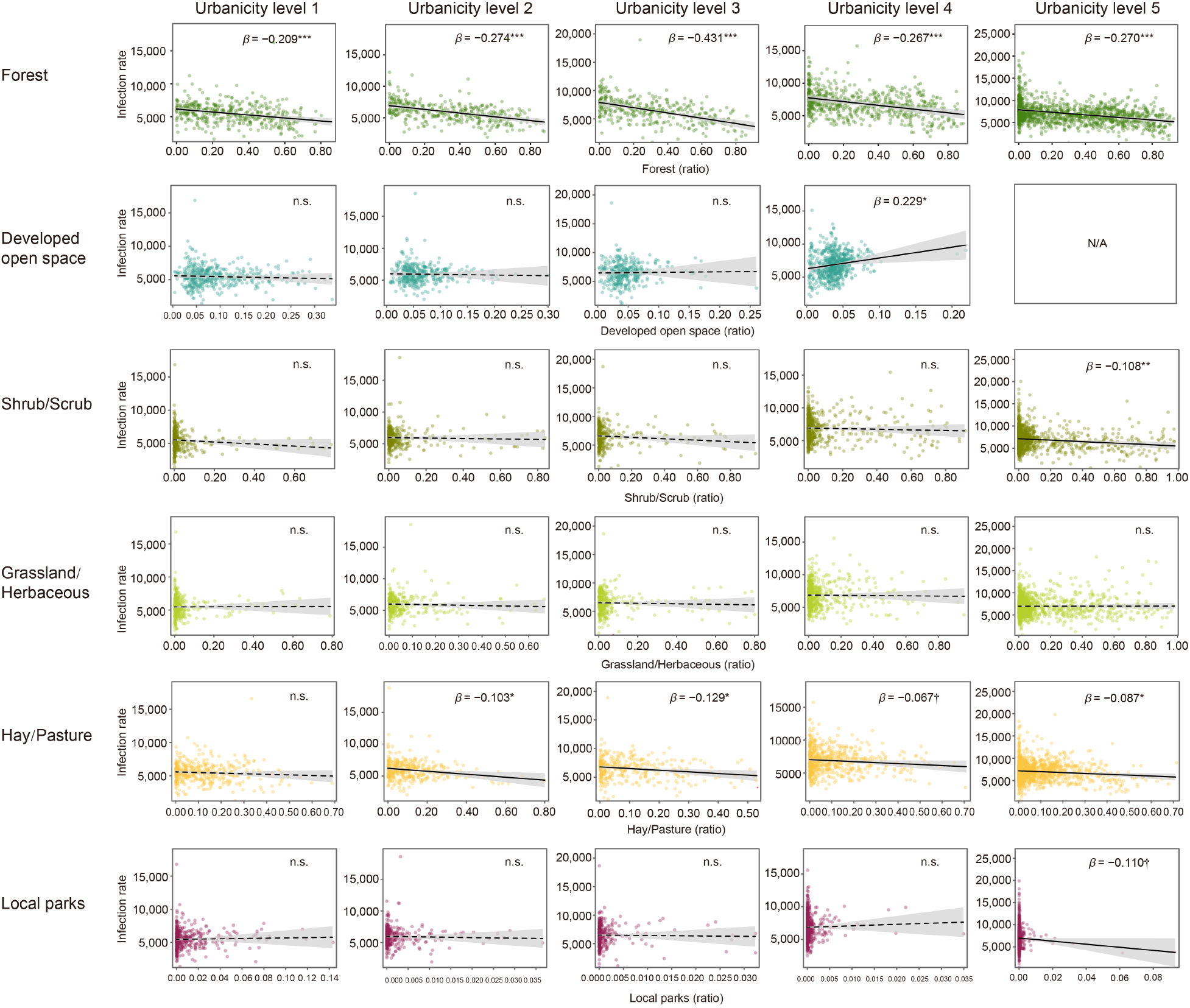
The association of six types of green spaces with SARS-CoV-2 infection rates across five levels of urbanicity. Each green space factor is calculated by dividing green space area by the total area of the county. Shaded areas represent the pointwise 95% confidence intervals. Points represent partial residuals. Lines represent the linear trend lines. A solid line indicates a statistically significant relationship, whereas a dashed line indicates a non-significant relationship. Note: ^ⴕ^ *p* < 0.05; * *p* < 0.01; ** *p* < 0.001; *** *p* < 0.0001.

### 3.3. Green space–infection rate associations across time periods

Next, we fitted five regression models to five time periods to explore temporal variations in green space–infection associations (Table S5). The additional explanatory power (Δ*R^2^*), resulting from the including green space factors, is significant across all time periods, and gradually increases from period 1 to period 5 (from 0.3% to 5.0%). Fig. 6 shows the individual effects of six different types of green space on SARS-CoV-2 infection rates for each the five time periods. Note that forest is significantly negatively associated with infection rates across all periods, except period 1, with the effect size increasing from period 2 to 5.

**Fig. 6.**
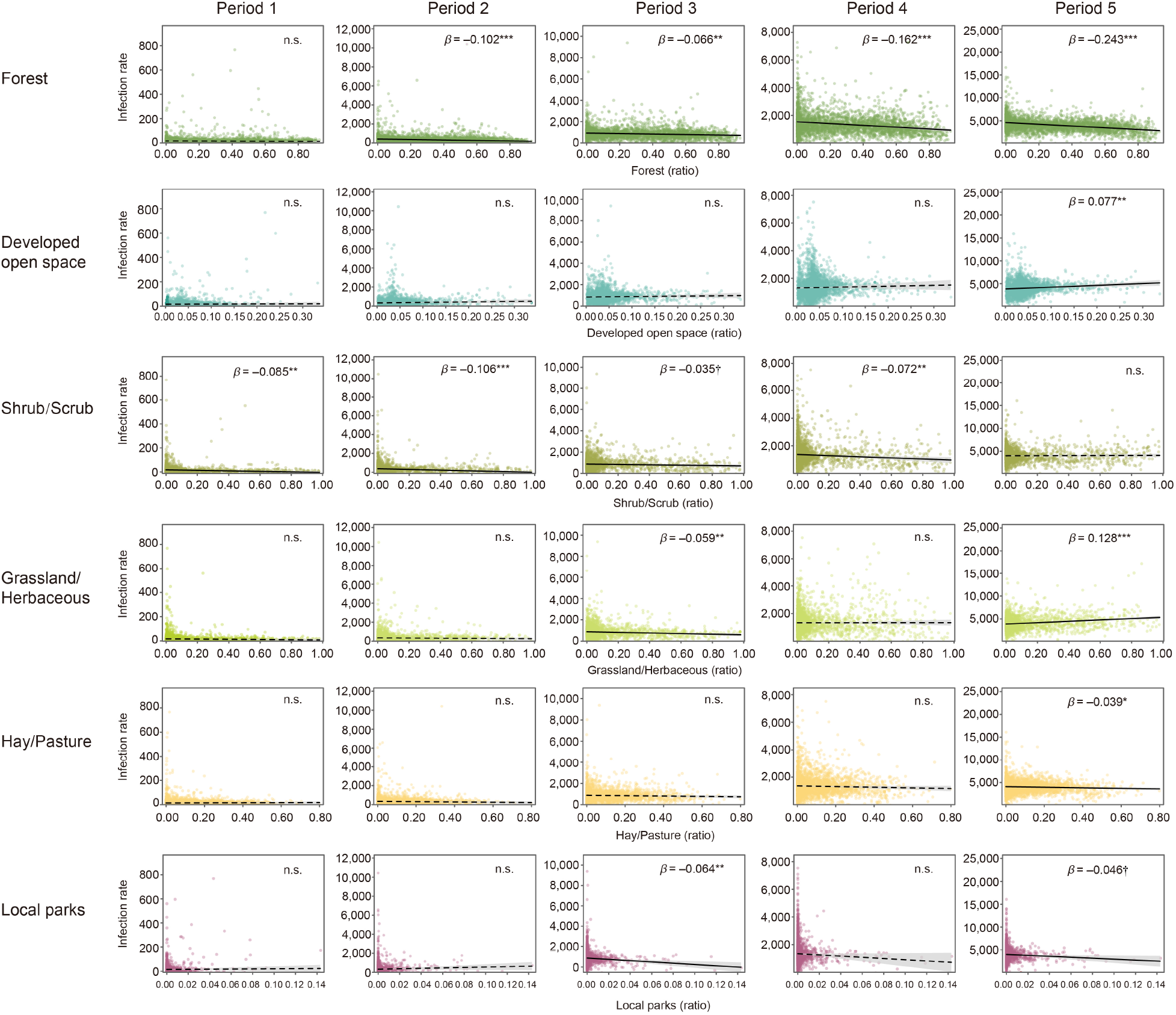
The effects of six types of green space on SARS-CoV-2 infection rates across five time periods. Each green space factor is calculated by dividing green space area by the total area of the county. Shaded areas represent pointwise 95% confidence intervals. Points represent partial residuals. Lines represent the linear trend line. Note: ^ⴕ^ *p* < 0.05; * *p* < 0.01; ** *p* < 0.001; *** *p* < 0.0001. Period 1: January 22 to March 30, 2020. Period 2: March 31 to June 7, 2020. Period 3: June 8 to August 15, 2020. Period 4: August 16 to October 23, 2020. Period 5: October 24 to December 31, 2020.

### 3.4. Forest–infection rate associations across five levels of urbanicity and five time periods

Given that forest has the most consistent and significant association with infection rates, we next examined the spatiotemporal patterns of the forest–infection rate association for 25 combinations of urbanicity levels and time periods. These associations, after controlling for all confounding factors, were presented in Table S6 and Fig. 7. Notice in Table S6 that the β values increases with successive periods across all urbanicity levels. Forest is significantly and negatively associated with infection rates at nearly all urbanicity levels in periods 4 and 5. Here again, the association of forest with infection rates is greatest at Urbanicity level 3 (*β* = −0.256 and −0.244 at time periods 4 and 5, respectively).

**Fig. 7.**
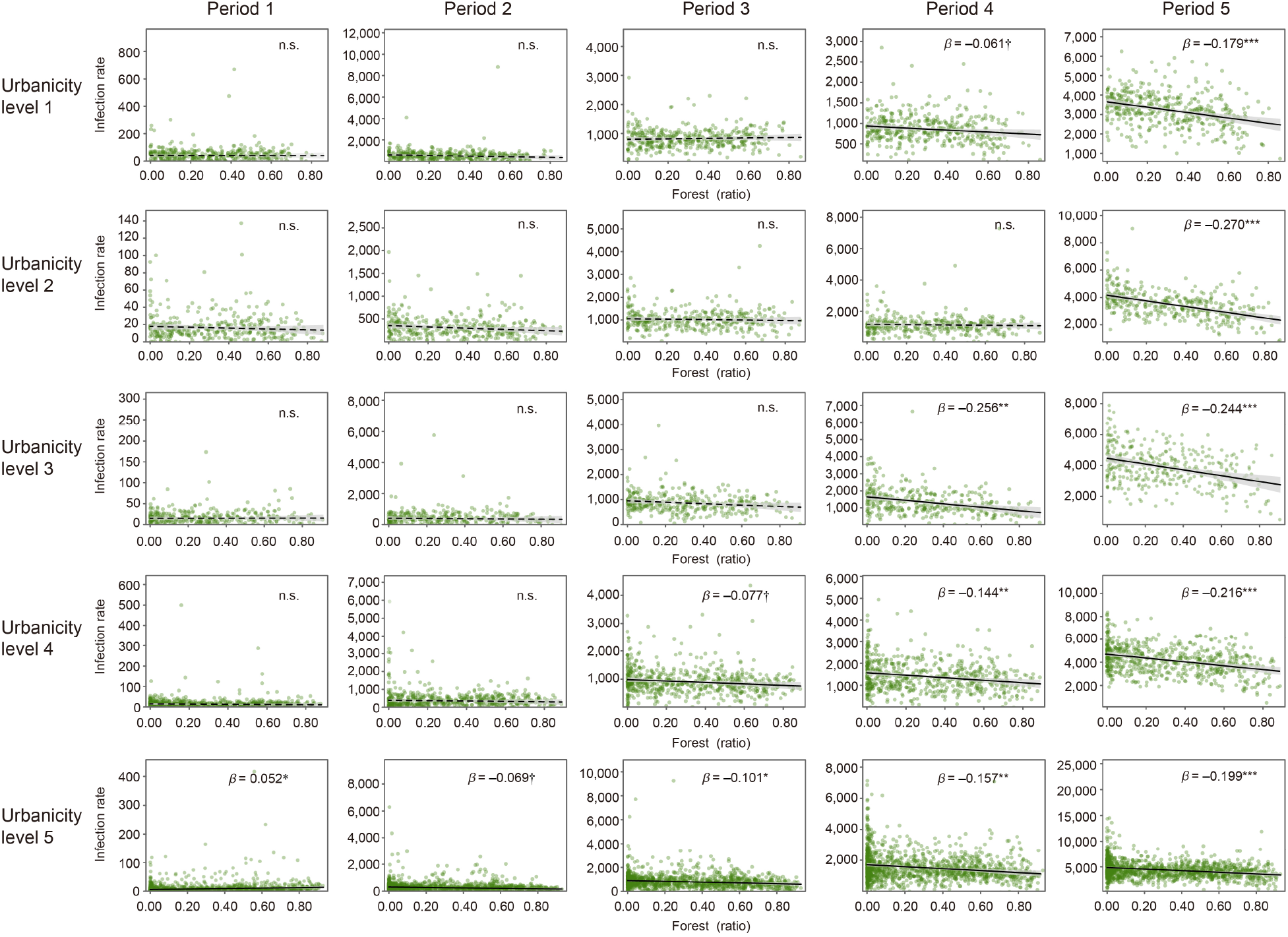
The spatiotemporal effects of forests on SARS-CoV-2 infection rates across five levels of urbanicity and five time periods. Shaded areas represent the pointwise 95% confidence intervals. Points represent partial residuals. Lines represent the linear trend lines. Note: ^ⴕ^ *p*< 0.05; * *p* < 0.01; ** *p* < 0.001; *** *p* < 0.0001.

### 3.5. Associations of population-weighted forest at varying distances with infection rates

Although these results revealed that forest has a negative association with infection rates at the county scale, the impact of the distance between forest and human settlements on this association is unclear. Thus, we now examined the association of population-weighted forest at various buffer distances (from 100 m to 50 km) with infection rates (Fig. 8).

**Fig. 8.**
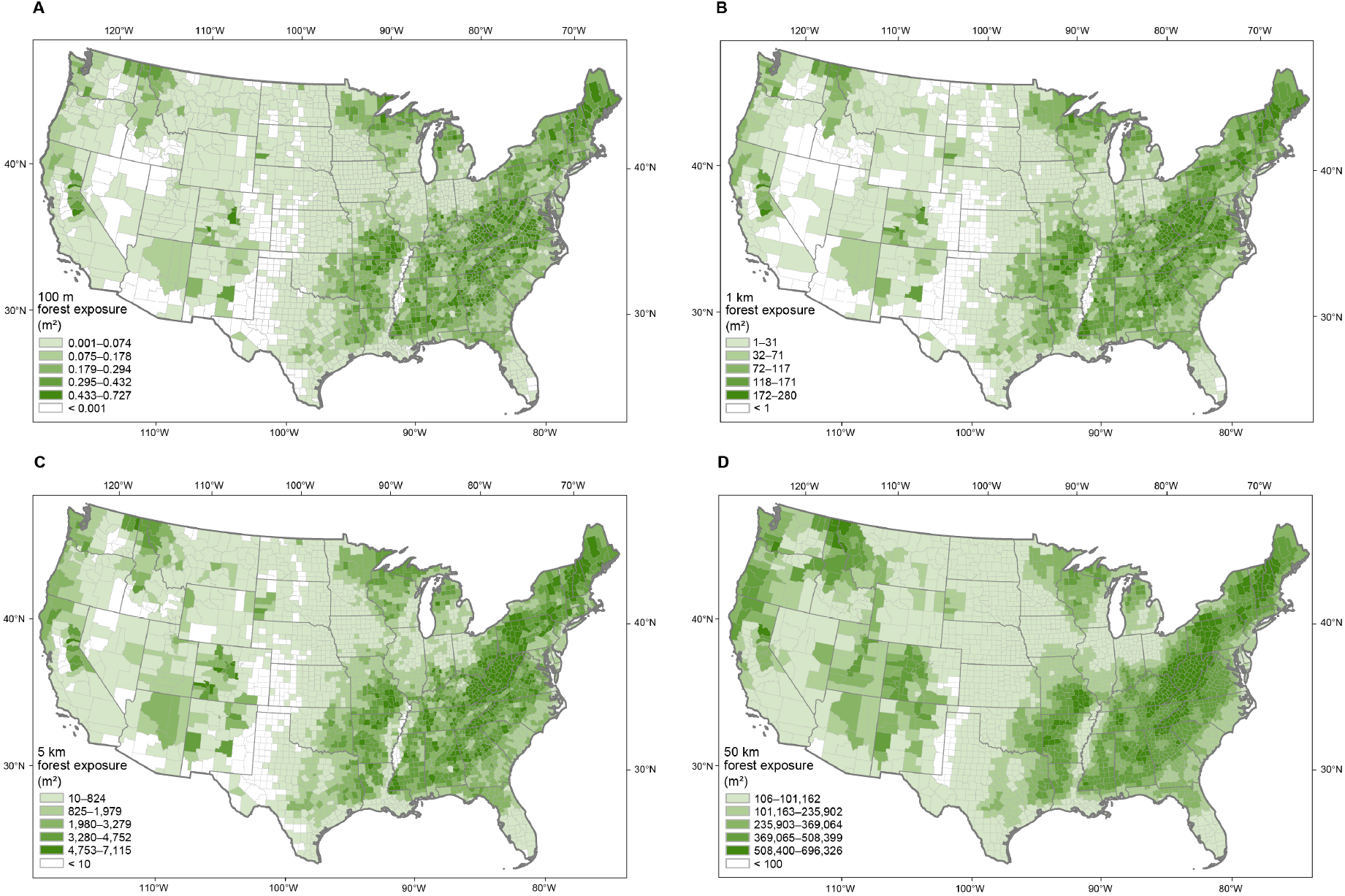
Population-weighted forest exposure at varying buffer distances at the county level. Population-weighted exposure to the nearest forest within a (A) 100-m, (B) 1-km, (C) 5-km, or (D) 50-km buffer distance at the county level. All population-weighted forest exposure values were calculated using Google Earth Engine.

First, we examined the relationship between forest within walking distance (0–5 km) with infection rates. This 5-km buffer distance is regarded as the longest reasonable walking distance for people in the United States (Yang & Diez-Roux, 2012). The effect size (*β*) and explanatory power (adjusted *R*^2^) of population-weighted forest exposures for various buffer distances were presented in Fig 8. The curves suggest a pattern of diminishing marginal effects. As Table S7 shows, the closest forest (within 100 m) contributed significantly to the overall effects seen for forest (*β* values: −0.250 vs −0.305, 82.0%; adjusted Δ*R*^2^ by forest: 0.029 vs 0.041, 70.7%).

With increasing buffer distance, the beneficial effects of forest exposure increase, however the extent of these increases gradually attenuates. At moderate buffer distances (≤ 1.0–1.4 km or 12.5–17.5 min given an average walking speed of 80 m/min), the effects of forest exposure reach a plateau, whereby further increases in buffer distance did not lead to further significant beneficial associations (Fig. 9 and Table S7). Compared to overall forest exposure, exposure to forest within a 1.0–1.4-km buffer distance is associated with the greatest reduction in infection rates within the optimal 5 km walking radius (e.g., for 1.0 km: *β* values: −0.273 vs −0.305, 89.5%; adjusted *R^2^* change by forest: 0.033 vs 0.041, 80.4%). This optimal walking distance is within the preferred maximum walking distance in the United States (Yang & Diez-Roux, 2012).

**Fig. 9.**
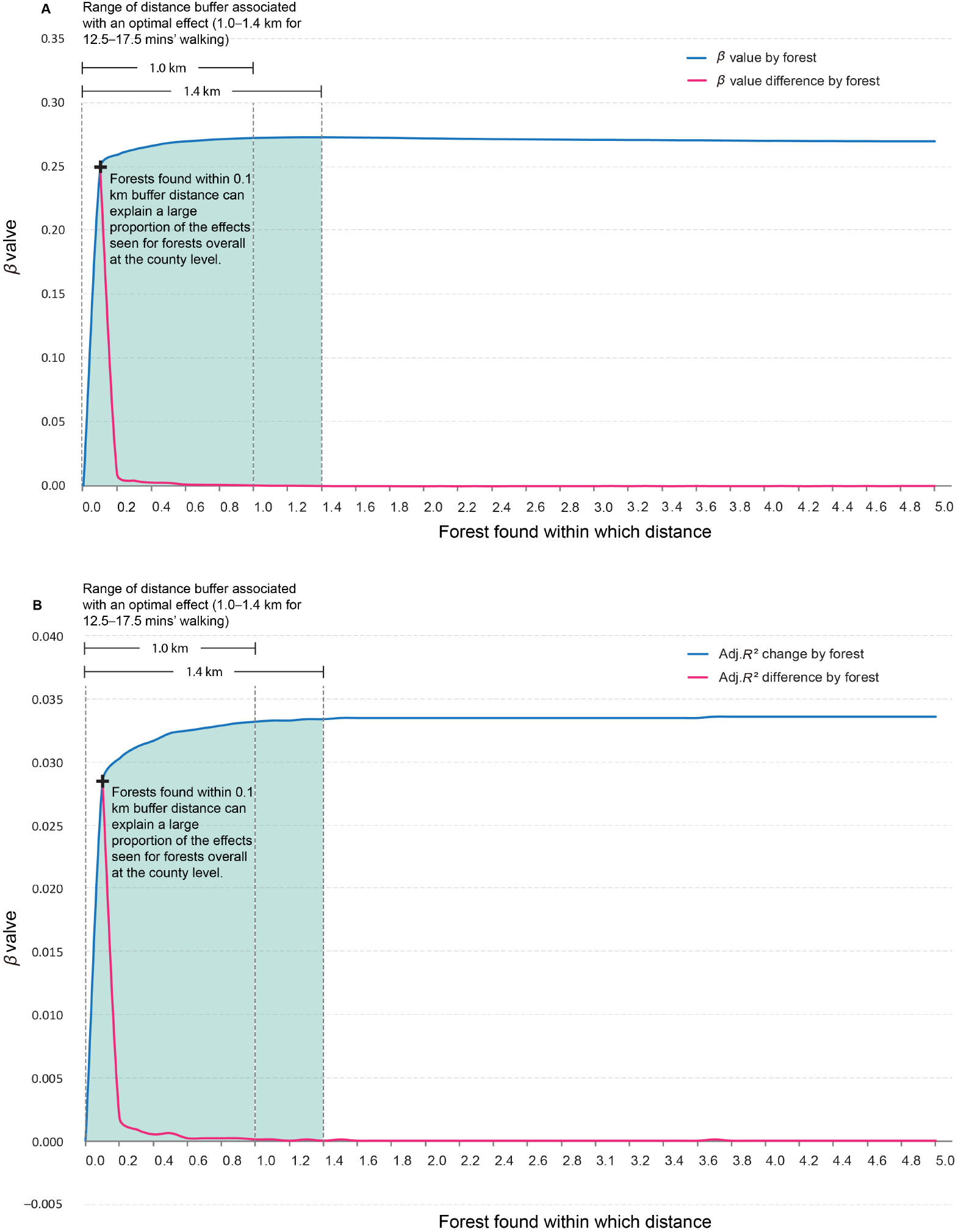
The effects of forest within walking distance (0–5 km) on infection rates. The effects are indicated by the *β* values and adjusted *R*^2^ values explained by forest in the regression models. The (A) *β* and (B) adjusted *R*^2^ values for forest exposure (blue line) continue to increase with increasing distances, however the size of the increments (shown in pink line) plateau with increasing distance.

We next investigated the association between forest beyond walking distance (5–50 km) and infection rate. The growth of *β* and incremental *R*^2^ remained small and stable as the buffer distance increases (Fig. 10 and Table S7). The curves for both *β* and incremental *R*^2^ plateau with a slight positive slope. Nevertheless, compared to forest falling within the 5-km walking range, we found that forest further afield is also related to infection rates, although the association is relatively minor. Using 50-km and 1.0-km distances for comparison, *β* values are −0.290 and −0.273, respectively, suggesting a 6.2% increase in the effect. Similarly, the adjusted *R*^2^ values are 0.036 and 0.033, respectively, suggesting a 9.1% increase in the effect. The proportion of the changes in *β* and adjusted *R*^2^ values accounted for by forest within 50 km relative to all forest in the county were 95.1% and 87.8%, respectively.

**Fig. 10.**
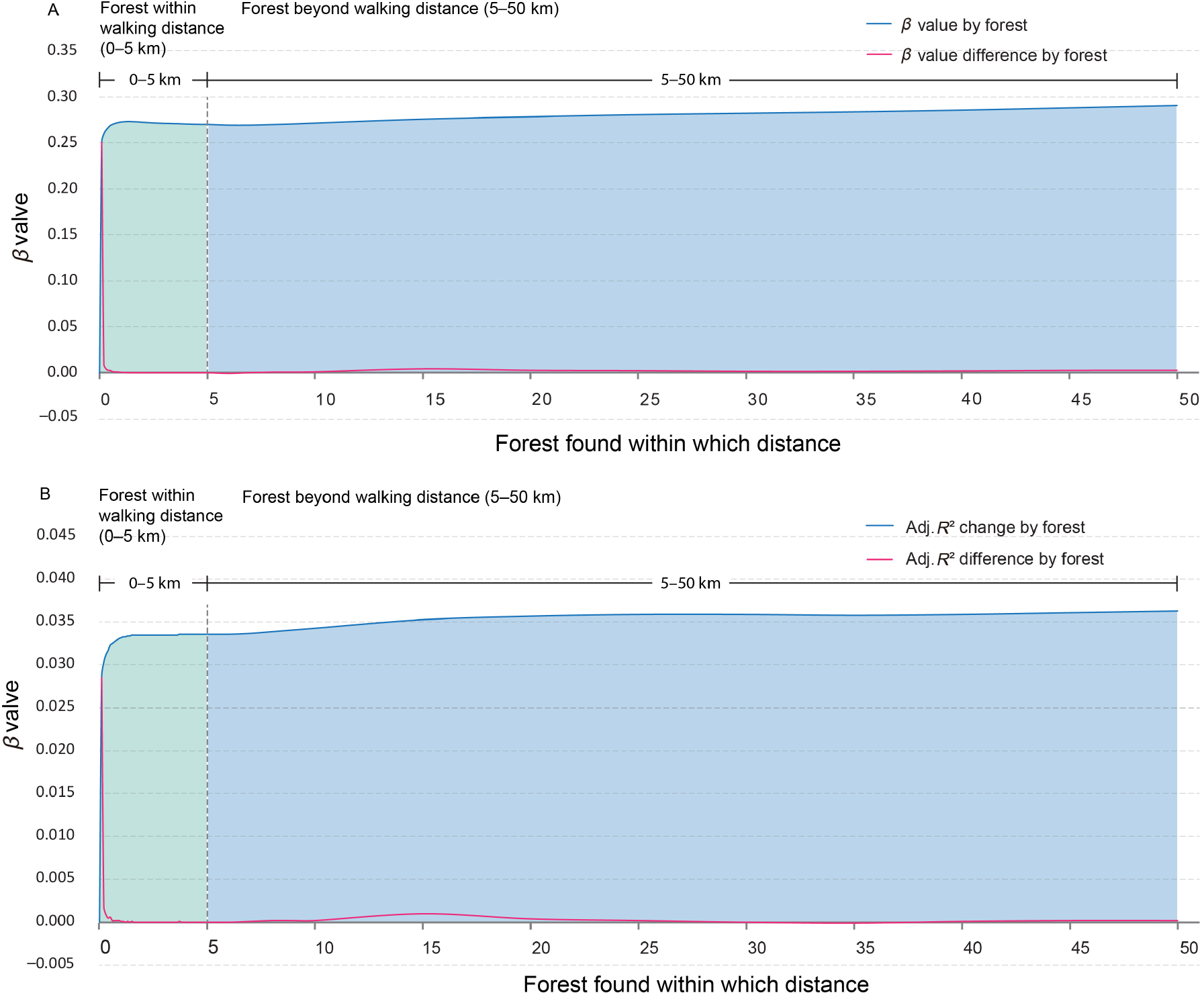
The effects of forest at varying buffer distances on infection rates (0–5 km and 5–50 km). Effects were determined by the *β* values and adjusted *R^2^* values explained by forest in the regression models. For both *β* and adjusted *R^2^* values, the portion of curves beyond walking distance (5–50 km) plateaued, with a slight positive slope. Nevertheless, compared to forest within the 5-km walking range, the forest further away could also reduce infection rates, although the effect was relatively minor.

## 4. Discussion

This study examined the association between green spaces and one-year of SARS-CoV-2 infection rates across all 3,108 counties in the contiguous United States after controlling for multiple categories of confounding factors. There were five major findings : (1) collectively, green spaces have an inverse association with SARS-CoV-2 infection rates; (2) forest has a stronger negative association with infection rates than all other types of green spaces; (3) the association between forest and infection rates is strongest for counties with a moderate level of urbanicity although the association remain significant for all five levels of urbanicity; (4) the association between forest and infection rates became stronger over the five time periods examined, and was strongest for the most recent time period when infection rates were at their highest; and (5) these associations can be largely explained by the presence of nearby forest within a moderate walking distance (≤ 1.0–1.4 km or about 12.5–17.5 minutes of walking on average).

In the following sections, we proposed key mechanisms to explain these main findings. We also discussed the novel contributions and implications of our findings. Finally, we discussed the limitations of this study and considered opportunities for future research.

### 4.1. How might green spaces reduce SARS-CoV-2 infection rates?

Before discussing potential mechanisms for reduced infection rates, it is important to note that the primary pathway of SARS-CoV-2 transmission is via aerosol particles (Klompas et al., 2020; Zhang, Li, Zhang, Wang, & Molina, 2020). Thus, people are primarily infected with SARS-CoV-2 by inhaling virus-containing droplets and aerosols exhaled by a human host (Bourouiba, 2020). Thus, compared to being indoors, being outdoors can reduce the risk of transmitting SARS-CoV-2 in two ways. First, outdoor environments often provide much stronger natural air movement, significantly reducing virus concentrations compared to indoor settings. In fact, a recent review suggested that the majority of SARS-CoV-2 infection clusters occurred in indoor settings (Leclerc et al., 2020). Second, outdoor environments, including large areas of green space, can support safer physical distancing, which reduces infection risks (Leclerc et al., 2020).

We found that, in general, green spaces were significantly inversely associated with SARS-CoV- 2 infection rates, even after controlling for confounding factors. Although regression models can only reveal associations, rather than causal relationships, we propose five mechanisms by which these effects could be explained.

#### 4.1.1. Mechanisms underlying reduced infection rates

##### (1) Encouraging outdoor activities

Green spaces, especially those with sufficient recreational opportunities and supporting facilities, can be regarded as a “socio-pedal”, in that they encourage people to leave indoor environments and participate in outdoor activities (Lu, Chen, et al., 2021; Lu, Zhao, et al., 2021; Schipperijn et al., 2013). One study reported that public spaces with more green landscapes can attract 90% more users than architecturally similar but barren public spaces (Sullivan et al., 2004). Similar findings have been reported by other studies (Chen et al., 2016; Liu et al., 2016). Social interaction is an essential aspect of human nature that can be suppressed for a short time, but cannot be sustained for long periods of time. Inevitably, despite restrictions on movement and socializing during the COVID-19 pandemic, people are likely to have had social gatherings in either indoor or outdoor settings (Geng et al., 2021). Green spaces not only invite people visit the outdoors more frequently (Coley, et al., 1997), but also seem to encourage them stay outdoors longer, reducing their time spent indoors (Braubach et al., 2017; Coley et al., 1997; Grahn & Stigsdotter, 2003).

##### (2) Promoting physical activities

Previous studies have reported that green spaces can promote physical activities, both prior to and during the COVID-19 pandemic (Cohen et al., 2007; Lu et al., 2019; Lu, Zhao, et al., 2021). Moreover, exercising while viewing green landscape can promote synergic health outcomes, which are better than those yielded by conducting exercise while lacking a green view (Pretty et al., 2005). Numerous studies have found that access to green spaces can reduce the risk of obesity, and individuals with obesity have been found to be more vulnerable to SARS-CoV-2 infection (Jia et al., 2021; Jordan & Adab, 2020; Sattar et al., 2020). Lastly, studies have suggested that physical activities in green spaces can enhance general health and immune function (Li et al., 2010), which may promote resistance to SARS-CoV-2 infection.

##### (3) Promoting mental restoration

Viewing or being in green environments can promote mental restoration by reducing stress (Hunter et al., 2019; Jiang et al., 2016; Ulrich et al., 1991), mental fatigue (Jiang et al., 2018; Kaplan, 1995; Sullivan & Li, 2021; Yu et al., 2018), depression (Min et al., 2017), and negative moods (Jiang et al., 2020). These mental health benefits are often interlinked with physical health benefits (Berman et al., 2008; Kaplan, 1995; Zhang, Yu, Zhao, Sun, & Vejre, 2020). Kuo (2015) argued that green spaces can promote immune function through promoting mental health, consistent with the findings of many other studies (e.g., Cohen et al., 2001; Segerstrom & Miller, 2004). Thus, the mental health benefits of green spaces can lead to improved physiological resistance to SARS-CoV-2 infection.

##### (4) Promoting social cohesion and cooperation

Social health benefits are often associated with the mental and physical health benefits of green spaces (Jiang, Zhang, et al., 2015; Sullivan et al., 2014). Studies have found that living beside green spaces leads to residents having less impulsiveness and aggressiveness, greater self-discipline, and more social connections with other residents than counterpart individuals living in barren areas (Branas et al., 2018; Kuo & Sullivan, 2001; Lee et al., 2019). Additionally, green spaces can reduce health disparities among populations with different socioeconomic or demographic statuses (Jiang et al., 2017; Mitchell & Popham, 2008). For example, a study found that a greater supply of green spaces was associated with a lower racial disparity in SARS-CoV-2 infection rates (Lu, Chen, et al., 2021). Social health benefits may lead to enhanced social security, cohesion, trust, and collaboration (Holtan et al., 2014; Jiang et al., 2017; Mazumdar et al., 2017), which are critical to reduce infection risk from a population behavior perspective, e.g., following orders to wear masks, maintaining physical distance, and reducing social gathering.

##### (5) Removing ambient pollutants

Green spaces, especially forest and trees, are critical for improving air quality. This is achieved through increased deposition rates of particulate matter on plant surfaces and/or the absorption of gaseous pollutants through leaf stomata (Janhäll, 2015; Kumar et al., 2019; Nowak et al., 2006; Nowak et al., 2014; Nowak et al., 2013; Nowak et al., 2018). These effects are not limited to forest found in urban areas, as forest in rural areas has been reported to also contribute to human health (Nowak et al., 2014). By inhibiting two major transmission routes of SARS-CoV-2, i.e., particles and aerosols, green spaces can reduce SARS-CoV-2 infection risk (Bourouiba, 2020). Another reason for this effect may be related to the expression of angiotensin-converting enzyme 2 (ACE-2) in the respiratory tract, a known receptor for coronaviruses. ACE-2 is overexpressed in individuals who are exposed ambient pollutants on a daily basis, especially nitrogen dioxide (NO_2_) and 2.5-µm– and 10-µm–diameter particulate matter (PM_2.5_ and PM_10_) (Fattorini & Regoli, 2020; Paital & Agrawal, 2020; Zhu et al., 2020).

#### 4.1.2. Why forest has a stronger negative association with infection rates than other green spaces?

In this study, a “forest” was defined as “a square (30 m × 30 m) dominated by trees, generally greater than 5 m tall, and accounting for greater than 20% of total vegetation cover” (MRLC consortium, 2019). Thus, “forest” does not only include large forest in the rural areas, but also includes areas with small patches of trees in urban, suburban, and rural areas (Fig. 2). We propose three reasons why forest may outperform other green spaces in reducing SARS-CoV-2 infection rates.

##### (1) Forest might be a stronger stimulus for outdoor activities compared to other green spaces

First, forest provides more possibilities for keeping safe physical distances than other green spaces, especially compared to other green spaces in urban areas. One study provided evidence for this argument (Lu, Zhao, et al., 2021): during the COVID-19 pandemic, more people chose to visit natural forest parks than urban parks for recreational and exercise purposes. An important explanation of this phenomenon is that compared to urban parks with limited open space, forest parks give people considerably more space to maintain a physical distance from other visitors. Second, experimental studies have found that tree canopies can create a more comfortable microclimate (Li et al., 2019; Ziter et al., 2019). Thus, forest may support outdoor activities more than other green spaces. Third, in this study, forest included lawn or grassland partially covered by tree canopies, which provide a more comfortable environment for outdoor activities than a lawn without shade or dense forest setting that has considerable shade.

##### (2) Forest might be more effective in boosting immune function

Forest has been reported to boost immune function in multiple ways; however, there is less evidence that other types of green spaces can do so. Several studies have demonstrated a beneficial effect of forest therapy on human’s weakened immune function (Li, 2010; Li et al., 2007; Lyu et al., 2019). These effects might be due to plant-derived phytoncides, which are antimicrobial volatile organic compounds that have been shown to reduce blood pressure, alter autonomic activity, and boost immune function by increasing the concentrations and activity of natural killer (NK) cells, among other effects (Komori et al., 1995; Li et al., 2006).

Moreover, although no study has determined whether enhanced immune function can directly reduce the risk of SARS-CoV-2 infection, studies have reported that patients with moderate-to-severe disease symptoms tend to have low concentrations of circulating NK cells, which is an indicator of impaired immune function (Jiang et al., 2020; Wang et al., 2020; Wilk et al., 2020). Additionally, reduced NK cell numbers have also been observed during the acute phase of SARS-CoV-2 infection (National Research Project for SARS, 2004). This evidence implies that forest reduces the risk and severity of SARS-CoV-2 infection by boosting NK cell populations.

##### (3) Forest has the potential to be a more mentally restorative green space than other green spaces

Forest has a complex three-dimensional profile, including tree trunks and canopies, and a higher level of biodiversity and greenness compared to other green spaces with relatively short and less biodiverse vegetation (Brockerhoff et al, 2017; Trochta et al, 2017). Thus, forest may have more restorative effects on mental fatigue and stress (Jiang et al., 2020). An experimental study found that streetscapes including trees more significantly reduced stress than those including grass or shrubs (Jiang, Larsen, et al., 2015; Jiang et al., 2016). Many other studies have also reported that visiting a forest park is more mentally restorative than visiting other green spaces, because of forest park’s higher ecological richness and greenness (e.g., Beil & Hanes, 2013; Wood et al., 2018).

##### (4) Forest may be more effective in removing ambient pollutants than other green spaces

Many studies have suggested that forest has a greater capacity for reducing air pollution. Forest can capture particulate pollutants more efficiently than grassland and shrubs, as forest often have more complex foliage, a larger number of trees, more diverse tree heights and canopy sizes, and more diverse tree species (Beckett et al., 2000). A nationwide study of trees and forest in the contiguous United States found that trees and forest removed approximately 17.4 million tonnes of air pollution in 2010, which was estimated to lead to 850 fewer deaths and 670,000 fewer incidents of acute respiratory symptoms (Nowak et al., 2006). One study conducted in Cambodia reported that deforestation was significantly associated with an increasing rate of acute respiratory infection in children (Pienkowski et al., 2017). Another study conducted in New York City revealed that increased tree density was significantly associated with a lower prevalence of early childhood asthma (Lovasi et al., 2008).

#### 4.1.3. Forest’ beneficial effect across urbanicity and time

First, the effect of forest on infection rates is most pronounced in moderately urbanized counties, although the effect remain significant across all five levels of urbanicity. As Table S8 shows, the residents of high-to-moderately high urbanized counties (urbanicity levels 1 and 2) had less forest exposure per capita compared to those in moderately urbanized counties (urbanicity level 3). Hence, green spaces in highly urbanized counties were shared by more people, making it more difficult for visitors to keep safe social distancing (Nobajas et al., 2020; Shoari et al., 2020). However, it is unclear why the forest–infection rate association was also weaker in rural counties (levels 4 and 5). It may be because forest in rural counties is likely to be private properties, which are not accessible to the public. Therefore, total land coverage may not accurately reflect the total green space available to the public, and thus the population infection rate might be less influenced by forest coverage.

Second, the effect of forest on infection rate was found to increase in strength as the pandemic progressed, and similar patterns were observed for other green spaces. There are two possible reasons for this. First, in the early period, the spread of SARS-CoV-2 might have been confined to hotspots and spread mainly via intimate social gatherings or institutional activities (such as family gatherings, party of close friends, or school activities) (Leclerc et al., 2020; Thakar, 2020). Residents of these local hotspots may have had different levels of green space exposure compared to the overall county-wide levels. However, in the later period of the pandemic, SARS-CoV-2 had spread widely across the whole county and affected most residents. Thus, residents’ exposure to green space may have been better reflected by county-level green space coverage in the late period, making the effect of green spaces more significant. Second, SARS-CoV-2 testing was highly inconsistent among counties and cases of infection were substantially underestimated in some areas during the early stage (Angulo et al., 2021), thus the numbers of confirmed cases of infection in the early periods are less reliable than those in the later periods.

#### 4.1.4. Forest within walking distance associated with reduce infection risk

A significant finding of this study was the identification of the optimal distance from forest for infection risk reduction, by considering the spatial distribution of populations across counties. The association between population-weighted forest exposure and SARS-CoV-2 infection rates followed the pattern of diminishing marginal utility. This finding suggests that it is critical to prioritize access to forest located within walking distance of critical masses of populations because doing so might significantly alleviate SARS-CoV-2 infection rates, in addition to providing other health benefits. The significant health benefits of nearby forest and other green spaces have been consistently reported (e.g., Corraliza & Collado, 2011; Cox et al., 2017; Lee et al., 2019; Oh et al., 2017). The construction and conservation of nearby forest is particularly valuable when a county has limited financial and human resources, due to the potential alleviation of burdens of healthcare systems. Additionally, forest beyond walking distance also appears to alleviate infection risks, potentially by reducing ambient pollutants at a regional scale (Escobedo & Nowak, 2009; Nowak et al., 2018).

### 4.2. Originality, significance, and contributions to knowledge and practice

This study makes original contributions to knowledge and practice that may influence future research, policymaking, planning, and design.

First, the spatiotemporal aspects of this study significantly enhance the validity and generalizability of its findings. The nationwide nature of the study provided an opportunity to investigate counties with different levels of urbanicity. The one-year span of the study also provided an opportunity to investigate the relatively complete course of a pandemic that developed from a low to high infection rate (Fig. 1). We considered the potential influence of various levels of urbanicity and time periods on the association of forest and other green spaces with reduced SARS-CoV-2 infection rates. The assumption of stationarity in relationships between environmental factors and health outcomes has been heavily criticized (Kwan, 2021). This study found that associations among green spaces and infection rates varied in significance and extent at different levels of urbanicity and over different time periods. These findings invite future studies that probe each outcome in greater depth so that we might better understand the associations identified here.

Second, this study identified a significant and strong negative association between green spaces and infection rates after controlling for socioeconomic and demographic, political, healthcare and testing, behavioral, and climate and environment factors. Such strong control for political and behavioral factors, such as presidential elections and the number of essential workers, has been rare in previous studies. These controls significantly increase the reliability of our findings.

Third, this study transcends a critical limitation of many ecological studies. Identifying a general positive or negative association between green spaces and health outcomes is inadequate to reveal the relationship and guide environmental interventions (Klompmaker et al., 2021). However, by determining the spatial relationship between forest and human settlements, we have greater confidence to propose an optimal buffer distance for forest that may achieve to maximal forest-associated reductions in infection rates.

### 4.3. Limitations and future research opportunities

There are four major opportunities that can be explored in future research following this study. This study reported correlational rather than casual findings. Still, the notion that exposure to green spaces, and forest in particular, lower infection rates from SARS-CoV-2 is plausible, based on the mechanisms proposed and a wealth of previous research. We suggest more experimental studies, including laboratory or natural experiments, be conducted to fully confirm these casual relationships (Jiang et al., 2021; Tyrvainen et al., 2014).

Second, we have restricted our interpretations on the significant findings to green spaces, and especially to landscapes that contain forest. We note, however, that many other factors are also significantly associated with infection rates, such as the Gini index, overcrowded housing, PM_2.5_ and PM_10_ concentrations, numbers of essential workers, modes of transport used for commuting, and public mask mandates. These factors all have significant potential to be the focal point of future studies.

Third, we did not use data collected in 2021, as vaccination programs were implemented in the majority of counties in the early months of 2021, and would have likely confound the relationship between green spaces and infection rates (BBC, 2020). Here again, we see this as an important opportunity for future research, as it is necessary to understand the extent to which varying levels vaccination rates alter the relationship between the supply of green spaces and SARS-CoV-2 infection rates and mortality rates.

Finally, although this study employed an ecological design that included a series of population-weighted assessments, the design is still vulnerable to the risk of ecological fallacy. That is, we should not draw inferences about individual levels of infection based on aggregate data gathered at the county-level. The fact that personal data related to individuals who were infected by SARS-COV-2 were not available makes overcoming this risk a challenge. Perhaps in some countries, where more data is available at the individual level, scientist can address this limitation.

## 5. Conclusion

This nationwide study is one initial study to investigate the year-long relationships between green spaces and SARS-CoV-2 infection rates at the county level. We found that forests and other green spaces have significant negative associations with the SARS-CoV-2 infection rates. Thus, preserving and developing forests and other green spaces might be important to alleviate the infection risk. Moreover, it is critical to preserve and develop forests within walking distance (≤ 1.0–1.4 km for 12.5–17.5 mins’ walking) of populations, to optimize the effects of forests on SARS-CoV-2 infection risks and maximize other potential health benefits for people.

## Contributions

BJ and YL proposed the research idea. YL and BJ conducted research design and guided the data collection and data analysis. BJ, YL, YWY, and CL conducted writing of manuscript. YWY, LC, XYW, XML, and JJW conducted the data collection and data treatment. LC and YWY conducted the data analysis with support from XYW and XML. BC and CW guided a key part of research design and data analysis. WCS reviewed and revised key parts of the manuscript. XML and XYW conducted the graphic design.

## Data Availability

1. landcover data of developed open space, forest, shrub/scrub, grassland/herbaceous, hay/pasture, and cultivated crops are extracted from National Land Cover Datasets in 2016.
2. Population-weighted forest exposure data is extracted from National Land Cover Datasets in 2016.
3. Local parks data is extracted from the Esri USA Parks dataset.
4. Tree canopy data is extracted from Tree Canopy Cover Datasets of the United States Forest Service (USFS).
5. NDVI data is extracted from NASA Moderate Resolution Imaging Spectroradiometer, which was calculated on Google Earth Engine.
6. SARS-CoV-2 Infection data is extracted from a dataset of the Centers for Disease Control and Prevention (CDC).
7. Socioeconomic and demographic data is extracted from a dataset of the US Census Bureau.
8. Healthcare and testing data is extracted from the CDC dataset.
9. Pre-existing chronic disease data is extracted from the CDC dataset.
10. Policy and regulation data is extracted from GitHub, CDC dataset, The National Governors Association dataset.
11. Behavioral factor data is extracted from the dataset of Behavioral Risk Factor Surveillance System (BRFSS) (County Health Rankings and Roadmaps)
12. Environmental factors data is extracted from the dataset of the National Land Cover Database (NLCD), US Census Bureau, and USGS National, Transportation Dataset (NTD) Airports, etc.

https://www.mrlc.gov/data/nlcd-2016-land-cover-conus

https://www.arcgis.com/home/item.html?id=578968f975774d3fab79fe56c8c90941

https://www.mrlc.gov/data/type/tree-canopy

https://usafacts.org/visualizations/coronavirus-covid-19-spread-map/

https://nccd.cdc.gov/DHDSPAtlas/Default.aspx

https://theuscovidatlas.org/

https://github.com/tonmcg/US_County_Level_Election_Results_08-20

https://www.nga.org/governors/

https://ephtracking.cdc.gov/DataExplorer/

https://www.countyhealthrankings.org/app/arizona/2020/measure/factors/9/data

