## Supplemental files for "Green spaces, especially forest, linked to lower SARS-CoV-2 infection rates: A one-year nationwide study"

### Supplementary materials

**List of tables**

Table S1. The definition of green space factors.

Table S2. The definition for socioeconomic and demographic factors, pre-existing chronic diseases, government policy, behavioral, and built environment factors.

Table S3. Descriptive data for SARS-CoV-2 infection rates, socioeconomic and demographic, pre-existing chronic disease, government policy, behavioral, built environment, and green space factors in the USA’s 3108 counties.

Table S4. Regression results of association between green spaces and infection rate by five urbanicity level.

Table S5. Regression results of association between green spaces and infection rate by five time periods.

Table S6. Regression result of associations between the ratio of forest and infection rate across five time periods and urbanicity levels.

Table S7. Regression results of association between population-weighted forest exposure and infection rate of varying buffer size.

Table S8. Mean forest exposure per capita and population-weighted forest exposure by urbanicity level.

Table S9. Descriptive data for confounding variables by urbanicity level.

Table S10. Regression results of association between the ratio of developed open space and infection rate across five time periods and urbanicity levels.

Table S11. Regression results of association between the ratio of shrub/scrub and infection rate across five time periods and urbanicity levels.

Table S12. Regression results of association between the ratio of grassland/herbaceous and infection rate across five time periods and urbanicity levels.

Table S13. Regression results of association between the ratio of hay/pasture and infection rate across five time periods and urbanicity levels.

Table S14. Regression results of association between the ratio of local parks and infection rate across five time periods and urbanicity levels.

**List of figures**

Fig. S1. Effect of forest on the SARS-CoV-2 infection rate across five time periods and urbanicity levels.
Fig. S2. Effect of developed open space on the SARS-CoV-2 infection rate across five time periods and urbanicity levels.
Fig. S3. Effect of shrub/scrub on the SARS-CoV-2 infection rate across five time periods and urbanicity levels.
Fig. S4. Effect of grassland/herbaceous on the SARS-CoV-2 infection rate across five time periods and urbanicity levels.
Fig. S5. Effect of hay/pasture on the SARS-CoV-2 infection rate across five time periods and urbanicity levels.
Fig. S6. Effect of local parks on the SARS-CoV-2 infection rate across five time periods and urbanicity levels.

Table S1. **The definition of green space factors**

| Categories | Classes | Official definition | Source | Link |
|---|---|---|---|---|
| **Green space factors** | Developed open space | Areas with a mixture of some constructed materials, but mostly vegetation in the form of lawn grasses. Impervious surfaces account for less than 20% of total cover. These areas most commonly include large-lot single-family housing units, parks, golf courses, and vegetation planted in developed settings for recreation, erosion control, or aesthetic purposes. | National Land Cover Datasets in 2016 | https://www.mrlc.gov/data/nlcd-2016-land-cover-conus |
| | Forest | Areas dominated by trees generally greater than 5 meters tall, and greater than 20% of total vegetation cover, including deciduous, evergreen, and mixed forest. Deciduous are tree species (more than 75%) shed foliage simultaneously in response to seasonal change, evergreen are tree species (more than 75%) maintain their leaves all year, and mixed are tree species (more than 75%) neither deciduous nor evergreen species. | National Land Cover Datasets in 2016 | https://www.mrlc.gov/data/nlcd-2016-land-cover-conus |
| | Shrub / Scrub | Areas dominated by shrubs; less than 5 meters tall with shrub canopy typically greater than 20% of total vegetation. This class includes true shrubs, young trees in an early successional stage or trees stunted from environmental conditions. | National Land Cover Datasets in 2016 | https://www.mrlc.gov/data/nlcd-2016-land-cover-conus |
| | Grassland / Herbaceous | Areas dominated by graminoid or herbaceous vegetation, generally greater than 80% of total vegetation. These areas are not subject to intensive management such as tilling but can be utilized for | National Land Cover Datasets in 2016 | https://www.mrlc.gov/data/nlcd-2016-land-cover-conus |

| | | | |
|---|---|---|---|
| | grazing. | | |
| Hay / Pasture | Areas of grasses, legumes, or grass-legume mixtures planted for livestock grazing or the production of seed or hay crops, typically on a perennial cycle. Pasture/hay vegetation accounts for greater than 20% of total vegetation. | National Land Cover Datasets in 2016 | https://www.mrlc.gov/data/nlcd-2016-land-cover-conus |
| Cultivated crops | Areas used for the production of annual crops, such as corn, soybeans, vegetables, tobacco, and cotton, and also perennial woody crops such as orchards and vineyards. Crop vegetation accounts for greater than 20% of total vegetation. This class also includes all land being actively tilled. | National Land Cover Datasets in 2016 | https://www.mrlc.gov/data/nlcd-2016-land-cover-conus |
| Local parks | Area of parks, gardens, and forests within the United States at local levels / county area, 2021. | Esri USA Parks | https://www.arcgis.com/home/item.html?id=578968f975774d3fab79fe56c8c90941 |
| Tree canopy | Percent tree canopy estimates for each pixel across all land covers and types. | Tree Canopy Cover Datasets of the United States Forest Service (USFS) | https://www.mrlc.gov/data/type/tree-canopy |
| NDVI | Average NDVI, 2020 | NASA Moderate Resolution Imaging Spectroradiometer | https://code.earthengine.google.com/# |
| Population-weighted forest exposure | Spatially population-weighted forest exposure aggregated to the county level, 2020; the buffer size ranges from 100m to 50km. | National Land Cover Datasets 2016, USGS NLCD | https://www.mrlc.gov/data/nlcd-2016-land-cover-conus |

Note: All data are at the county level.

Table S2. **The definition for socioeconomic and demographic factors, pre-existing chronic diseases, government policy, behavioral, and built environment factors**

| Variable Categories | Variables | Item description (formula) | Official definition | Source | Link |
|---|---|---|---|---|---|
| **SARS-CoV-2 Infection rate** | Infection rate | Total cases per 100k | Daily county-level cumulative totals of positive cases from 01/22 to 12/31 in 2020 | Centers for Disease Control and Prevention (CDC), state- and local-level public health agencies. | https://usafacts.org/visualizations/coronavirus-covid-19-spread-map/ |
| | Infection rate period 1-5 | Cases per 100k for 5 periods | Daily county-level cumulative totals of positive cases 01/22-03/30, 03/31-06/07, 06/08-08/15, 08/16-10/23, 10/24-12/31 in 2020 | Centers for Disease Control and Prevention (CDC), state- and local-level public health agencies. | https://usafacts.org/visualizations/coronavirus-covid-19-spread-map/ |
| **Socioeconomic and demographic factors** | Population density | Total population / county area | County population, 2019 | US Census Bureau | https://nccd.cdc.gov/DHDSPAtlas/Default.aspx |
| | Female household ratio | Families with Female Head of Household / Total families | Percent of families with female head of household, 2014 to 2018 (5-year, average) | US Census Bureau | https://nccd.cdc.gov/DHDSPAtlas/Default.aspx |
| | Black or African American non-Hispanic | Black or African American non-Hispanic Population / Total population | Percent of Black or African American non-Hispanic Population 2014 to 2018 (5-year, average) | US Census Bureau | https://nccd.cdc.gov/DHDSPAtlas/Default.aspx |
| | White non-Hispanic | White non-Hispanic Population / Total population | Percent of white non-Hispanic Population 2014 to 2018 (5-year, average) | US Census Bureau | https://nccd.cdc.gov/DHDSPAtlas/Default.aspx |
| | Hispanic/Latino | Hispanic/Latino Population / Total | Percent of Hispanic/Latino Population 2014 to 2018 (5-year, average) | US Census Bureau | https://nccd.cdc.gov/DHDSPAtlas/Default.aspx |

| | | | | |
|---|---|---|---|---|
| | population | | | |
| Population aged 65 and older | Populations 65 years and over/ total population | Population aged 65 and older Percent 2014 to 2018 (5-year, average) | US Census Bureau | https://nccd.cdc.gov/DHDSPAtlas/Default.aspx |
| Median household income | | Income of the householder and all other individuals 15 years old and over in the household, 2018 | US Census Bureau | https://nccd.cdc.gov/DHDSPAtlas/Default.aspx |
| Gini Index | | The difference between a diagonal line (the purely proportionate distribution) and the distribution of actual values (a Lorenz curve), 2014-2018 (5-year, average) | US Census Bureau | https://nccd.cdc.gov/DHDSPAtlas/Default.aspx |
| Poverty rate | Population living in Poverty / Total population | Individuals in family that total income is less than the family's money threshold defined by Census Bureau are considered in poverty, 2018 | US Census Bureau | https://nccd.cdc.gov/DHDSPAtlas/Default.aspx |
| Median housing value | Median value of the housing value | Median value of owner-occupied housing units, 2014-2018 (5-year, average) | US Census Bureau | https://nccd.cdc.gov/DHDSPAtlas/Default.aspx |
| Food stamp supplement | Population with food stamp / Total population | Percentage food stamp/supplemental nutrition assistance program recipient, a federally funded program that "permit low-income households to obtain a more nutritious diet", 2016 | US Census Bureau | https://nccd.cdc.gov/DHDSPAtlas/Default.aspx |
| Unemployment rate | Number of unemployed people (Ages 16+) / Total civilian labor force | The unemployment rate represents the number of unemployed. people as a percentage of the civilian labor force, 2018 | US Census Bureau | https://nccd.cdc.gov/DHDSPAtlas/Default.aspx |
| Population without | Population without 4 plus | People whose highest degree was a high school diploma or its | US Census Bureau | https://nccd.cdc.gov/DHDSPAtlas/ |

| | | | | | |
|---|---|---|---|---|---|
| | college degree | Years College / Total population | equivalent and people who attended college but did not receive a degree, ages 25+, 2014-2018 (5-year, average) | | Default.aspx |
| | Population without a high school diploma | Population without a high school diploma/ Total population | People who drop out in high school or before high school and never attended high school, ages 25+, 2014-2018 (5-year, average) | US Census Bureau | https://nccd.cdc.gov/DHDSPAtlas/Default.aspx |
| **Healthcare and testing** | Testing rate | Testing rate per 100k | Number of Covid19 tests in county normalized to per 100k from 01/22 to 12/31 and by 5 infection time periods. | The US Covid Atlas, CDC | https://theuscovidatlas.org/ |
| | Health insurance coverage | Percent without insurance | Percent of population with no health insurance under age 65, 2017 | Deaths National Vital Statistics System. Bridged-Race Postcensal Population Estimates from National Center for Health Statistics. | https://nccd.cdc.gov/DHDSPAtlas/Default.aspx |
| **Pre-existing chronic disease factors** | Hypertension morality | Hypertension death rate per 100k | Hypertension death rate per 100k, 2016-2018 | Deaths National Vital Statistics System. Bridged-Race Postcensal Population Estimates from National Center for Health Statistics. | https://nccd.cdc.gov/DHDSPAtlas/Default.aspx |
| | Heart failure mortality | Heart failure death rate per 100k | Heart failure death rate per 100k, 2016-2018 | Deaths National Vital Statistics System. Bridged-Race Postcensal Population Estimates from National Center for Health Statistics. | https://nccd.cdc.gov/DHDSPAtlas/Default.aspx |
| | Stroke mortality | Stroke death rate per 100k | Stroke death rate per 100k, 2016-2018 | Deaths National Vital Statistics System. Bridged-Race Postcensal Population Estimates from National Center for Health Statistics. | https://nccd.cdc.gov/DHDSPAtlas/Default.aspx |
| | Diagnosed diabetes | Population with diagnosed | Percent of diagnosed diabetes (aged 20+), 2016 | CDC, Division of Diabetes | https://nccd.cdc.gov/DHDSPAtlas/Default.aspx |

| | | | | | |
|---|---|---|---|---|---|
| | | diabetes / Total population | | | |
| | Obesity | Population with obesity / Total population | Percent of obesity (aged 20+), 2016 | CDC, Division of Diabetes | https://nccd.cdc.gov/DHDSPAtlas/Default.aspx |
| **Policy and regulation** | Presidential vote rates | All electoral votes called for Biden / Total votes | Ratio of democrats electoral votes | GitHub collected 2020 election results published by Fox News, Politico, and the New York Times | https://github.com/tonmcg/US_County_Level_Election_Results_08-20 |
| | State Governor Party | | State government administrated party of year in 2020 (1=democratic party, 0=republican party). | The National Governors Association | https://www.nga.org/governors/ |
| | Stay-at-home orders | Sum of stay-at-order restriction level / days of restriction | State-issued stay-at-home order restriction level from 01/22 to 12/31 and by 5 infection time periods.<br>Restriction level:<br>(1) No order found,<br>(2) Advisory/Recommendation,<br>(3) Mandatory - at-risk in certain areas of state,<br>(4) Mandatory - minors only,<br>(5) Mandatory - at-risk people only,<br>(6) Mandatory - all people in certain areas of state,<br>(7) Mandatory - all people. | CDC obtained from state and territorial government websites | https://ephtracking.cdc.gov/DataExplorer/ |
| | Public mask mandates | Sum of public mask mandate restriction level/ days of restriction | U.S. state and territorial public mask mandates from 01/22 to 12/31 and by 5 infection time periods.<br>Restriction level:<br>(1) No restriction found,<br>(2) Authorized to fully reopen,<br>(3) Open with limitations,<br>(4) Curbside/delivery only, | CDC obtained from state and territorial government websites | https://ephtracking.cdc.gov/DataExplorer/ |

| | | | | | |
|---|---|---|---|---|---|
| | | | (5) Closed. | | |
| | Business closing and reopening orders | Sum of business closing and reopening order restriction level/ days of restriction | State issued bar and restaurant closing and reopening orders from 01/22 to 12/31 and by 5 infection time periods, Restriction level:<br>(1) No restriction found,<br>(2) Authorized to fully reopen,<br>(3) Open with limitations,<br>(4) Curbside/delivery only,<br>(5) Closed. | CDC obtained from state and territorial government websites | https://ephtracking.cdc.gov/DataExplorer/ |
| **Behavioral factor** | Smoker | | Proportion of current smokers in 2017 | Behavioral Risk Factor Surveillance System (BRFSS) (County Health Rankings and Roadmaps) | https://www.countyhealthrankings.org/app/arizona/2020/measure/factors/9/data |
| | Essential worker | | Percent of essential workers 2019, based on Chicago Metropolitan Agency for Planning classification (CMAP) | The US Covid Atlas, American Community Survey (ACS) | https://theuscovidatlas.org/ |
| | Mobility | | The median of the max-distance mobility for all samples in the specified region | Descartes Labs | https://seandavi.github.io/sars2pack/reference/descartes_mobility_data.html |
| | Mobility index | | The percent of normal m50 in the region, with normal mobility defined from 2020-02-17 to 2020-03-07 | Descartes Labs | https://seandavi.github.io/sars2pack/reference/descartes_mobility_data.html |
| | Point of interest visit (POI) | | Aggregated data of county-level foot traffic to different out-of-home activities from March 1, 2020 to March 21,2020 | SafeGraph | https://github.com/JieYingWu/COVID-19_US_County-level_Summaries/tree/master/data/out_of_home_activity |
| | Commute to work by public transportation | | Percent of workers over 16 years that used public transportation (excluding taxi cabs), 2012-2016 | American Community Survey (ACS) | https://ephtracking.cdc.gov/showCommunityDesignElements |
| | Commute to work by | | Percent of workers over 16 years that used car truck van, | American Community Survey (ACS) | https://ephtracking.cdc.gov/showCommunityDesignElements |

| | | | | | |
|---|---|---|---|---|---|
| | private vehicles | | 2012-2016 | | |
| | Commute to work by walk or bicycle | | Percent of workers over 16 years that used bicycle & walking, 2012-2016 | American Community Survey (ACS) | https://ephtracking.cdc.gov/showCommunityDesignElements |
| | Leisure-time physical inactivity | Population who lacks physical activity / Total population | Leisure Time Physical Inactivity 2016 Age-Adjusted Percentage; Adults Aged 20+ Years, 2016-2018 | CDC Division of Diabetes | https://nccd.cdc.gov/DHDSPAtlas/Default.aspx |
| **Environmental factors** | Developed intensity | Areas characterized by a high percentage (30 percent or greater) of constructed materials | "Developed, low intensity" is 20-49 percent impervious surface, "Developed, medium intensity" is 50-79 percent impervious surface, and "Developed, high intensity" is 80 percent or more impervious surface. | National Land Cover Database (NLCD) | https://ephtracking.cdc.gov/showCommunityCharacteristicsIndicators |
| | Severe housing problems | Rate of households living with severe housing problems | Percentage of occupied housing units with at least one of the following problems: lack of complete kitchen facilities, lack of plumbing facilities, overcrowding or severely cost-burdened occupants, 2010-2014 | US Census Bureau | https://nccd.cdc.gov/DHDSPAtlas/Default.aspx |
| | Crowded housing | | Percent of housing units with more people than rooms, 2016 | National Environmental Public Health Tracking Network | https://ephtracking.cdc.gov/showIndicatorPages.action?selectedContentAreaAbbreviation=22&selectedIndicatorId=110&selectedMeasureId= |
| | Proximity to highway | Percent of the population live within 150m to the highway | Proportion of population within census tracts (US Census 2010) within a 150m buffer around all major highways (freeways and expressways) calculated using Esri ArcGIS 10.1 and summed from the census tract level to the county level, 2010. | National Environmental Public Health Tracking Network | https://ephtracking.cdc.gov/showCommunityDesignElements |
| | Airport | Density of | Total number of airport / county | USGS National | https://www.sciencebase.gov/catalo |

| | | | | |
|---|---|---|---|---|
| density | airports | area, 2014 | Transportation Dataset (NTD) Airports | g/item/4f70b1f4e4b058caae3f8e16 |
| Railway density | Density of railway | Total railway length/county area, 2014 | USGS National Transportation Dataset (NTD) Railroad | https://www.sciencebase.gov/catalog/item/4f70b1f4e4b058caae3f8e16 |
| Highway and secondary road density | Density of highway and secondary road | total highway and secondary road length/county area, 2014 | USGS National Transportation Dataset (NTD) Highway and Secondary | https://www.sciencebase.gov/catalog/item/4f70b1f4e4b058caae3f8e16 |
| Sun exposure | Average sunlight exposure | The total amount of solar energy of all types received by a particular surface horizontal to the sun, 2012. | Centers for Disease Control and Prevention (CDC) | https://ephtracking.cdc.gov/DataExplorer/ |
| PM2.5 | PM2.5 annual average ambient concentration | Downscale (DS) modeled predictions for counties and days without monitoring data and using Air Quality System (AQS) data for counties and days with monitoring data, 2016. | Centers for Disease Control and Prevention (CDC) | https://ephtracking.cdc.gov/DataExplorer/ |
| PM10 | Mean PM10 Concentration in each county, 2020 | | United States Environmental Protection Agency (EPA) | https://www.epa.gov/outdoor-air-quality-data/download-daily-data |
| NO2 | Mean NO2 Concentration in each county, 2020 | | United States Environmental Protection Agency (EPA) | https://www.epa.gov/outdoor-air-quality-data/download-daily-data |
| Precipitation | Average monthly precipitation | State-level average monthly precipitation in inches from 01/22 to 12/31 and by 5 infection time periods. | U.S. Climate Divisional Database | https://www.ncdc.noaa.gov/cag/ |
| Maximum temperature | Average monthly maximum temperature | State-level average monthly maximum temperature in Fahrenheit from 01/22 to 12/31 in 2020 and by 5 infection time | U.S. Climate Divisional Database | https://www.ncdc.noaa.gov/cag/ |

| | | | | | |
|---|---|---|---|---|---|
| | | | periods. | | |
| | Minimum temperature | Average monthly minimum temperature | State-level average monthly minimum temperature in Fahrenheit from 01/22 to 12/31 in 2020 and by 5 infection time periods. | U.S. Climate Divisional Database | https://www.ncdc.noaa.gov/cag/ |
| | Average temperature | Average monthly average temperature | State-level average temperature in Fahrenheit from 01/22 to 12/31 in 2020 and by 5 infection time periods. | U.S. Climate Divisional Database | https://www.ncdc.noaa.gov/cag/ |
| | Relative humidity | Mean relative humidity in each county, 2020, | | National Oceanic and Atmospheric Administration (NOAA) | https://www.ncei.noaa.gov/data/global-hourly/archive/csv/ |
| | Wind speed | Average wind speed | Wind speed by a simulation model based on a dataset of the 5-minute time resolution, from 2007 to 2013. | The National Renewable Energy Laboratory (NREL) | https://www.ncei.noaa.gov/data/global-hourly/archive/csv/ |
| **Urbanicity levels** | Urbanicity level 1 to 5 | | Urbanicity level 1 include large central and fringe metro counties in metropolitan statistical areas (MSAs) with population of one million or more; level 2 includes medium metro counties in MSAs with population of 250,000 to 999,999; level 3 includes small metro counties in MSAs with population less than 250,000; level 4 includes counties in micropolitan statistical areas; level 5 includes counties that did not qualify as metropolitan or micropolitan. | National Center for Health Statistics | https://www.cdc.gov/nchs/data_access/urban_rural.htm |

Note: All data are at the county level.

Table S3. **Descriptive data for SARS-CoV-2 infection rates, socioeconomic and demographic, pre-existing chronic disease, policy and regulation, behavioral, built environment, and green space factors.**

| Categories | Variables | Min | Max | Mean | SD | Unit or Formula | VIF Test |
|---|---|---|---|---|---|---|---|
| **SARS-CoV-2 Infection rate** | Infection rate | 420.885 | 27173.734 | 6570.950 | 2762.664 | Cases per 100k | N/A |
| | Infection rate P1 | 0.000 | 964.232 | 15.553 | 43.675 | Cases per 100k | N/A |
| | Infection rate P2 | 0.000 | 12327.189 | 338.600 | 633.546 | Cases per 100k | N/A |
| | Infection rate P3 | 0.000 | 10995.013 | 858.663 | 848.936 | Cases per 100k | N/A |
| | Infection rate P4 | 0.000 | 8480.792 | 1334.447 | 1014.718 | Cases per 100k | N/A |
| | Infection rate P5 | 137.687 | 25771.325 | 4023.687 | 1973.626 | Cases per 100k | N/A |
| | Infection rate U1 | 1204.077 | 19133.286 | 5473.926 | 2007.042 | Cases per 100k | N/A |
| | Infection rate U2 | 1079.317 | 19831.301 | 5951.853 | 2048.693 | Cases per 100k | N/A |
| | Infection rate U3 | 843.124 | 21652.334 | 6494.541 | 2667.602 | Cases per 100k | N/A |
| | Infection rate U4 | 733.718 | 18853.211 | 6868.337 | 2785.377 | Cases per 100k | N/A |
| | Infection rate U5 | 420.885 | 27173.734 | 6986.236 | 3028.206 | Cases per 100k | N/A |
| **Socioeconomic and demographic factors** | Population density | 0.094 | 19625.842 | 98.221 | 559.972 | Persons per $km^2$ | Retained |
| | Female household ratio | 0.000 | 40.800 | 11.081 | 4.308 | Ratio | Removed |
| | Black or African American non-Hispanic | 0.000 | 87.400 | 8.957 | 14.520 | Ratio | Retained |
| | White non-Hispanic | 0.700 | 100.000 | 76.791 | 19.853 | Ratio | Removed |
| | Hispanic/Latino | 0.000 | 99.000 | 9.249 | 13.855 | Ratio | Removed |
| | Population aged 65 and older | 3.800 | 55.600 | 18.428 | 4.542 | Ratio | Retained |
| | Median household income | 25000.000 | 140000.000 | 52657.01 | 13825.13 | USD$ | Removed |
| | Gini index | 0.257 | 0.665 | 0.446 | 0.036 | Range 0-1 | Retained |
| | Poverty rate | 2.600 | 54.000 | 15.180 | 6.112 | Ratio | Removed |

| | | | | | | | |
|---|---|---|---|---|---|---|---|
| | Median housing value | 21000.00 | 1057000.00 | 146157.40 | 89047.61 | USD$ | Retained |
| | Food stamp supplement | 0.400 | 55.700 | 13.972 | 7.270 | Ratio | Removed |
| | Unemployment rate | 1.300 | 18.100 | 4.093 | 1.397 | Ratio | Retained |
| | Population without college degree | 21.500 | 100.000 | 78.452 | 9.436 | Ratio | Removed |
| | Population without high school diploma | 1.200 | 66.300 | 13.448 | 6.342 | Ratio | Retained |
| **Healthcare and testing** | Testing rate | 0.000 | 1711.099 | 170.197 | 107.693 | Per 100k | Retained |
| | Health insurance coverage | 2.300 | 33.700 | 11.418 | 5.106 | Ratio | Retained |
| **Pre-existing chronic disease factors** | Heart failure mortality | 19.400 | 304.300 | 108.314 | 25.488 | Per 100k | Retained |
| | Stroke mortality | 14.000 | 92.500 | 39.776 | 8.153 | Per 100k | Retained |
| | Hypertension morality | 19.300 | 587.300 | 131.825 | 55.366 | Per 100k | Retained |
| | Diagnosed diabetes rate | 2.200 | 28.700 | 10.506 | 3.526 | Ratio | Retained |
| | Obesity rate | 12.30 | 57.90 | 32.76 | 5.68 | Ratio | Retained |
| **Policy and regulation** | Presidential vote rates | 0.031 | 0.945 | 0.338 | 0.163 | Ratio | Removed |
| | State Governor Party | 0.000 | 1.000 | 0.435 | 0.496 | Democratic / Republican | Retained |
| | Stay-at-home orders | 3.856 | 6.897 | 4.441 | 0.447 | Range 1-7 | Retained |
| | Public mask mandates | 0.000 | 1.000 | 0.497 | 0.339 | Yes/No | Retained |
| | Business closing and reopening | 4.781 | 8.233 | 6.331 | 0.683 | Range 1-5 | Retained |
| **Behavioral factor** | Smoker | 5.909 | 41.491 | 17.446 | 3.554 | Ratio | Retained |
| | Essential worker | 0.178 | 0.791 | 0.526 | 0.073 | Ratio | Retained |
| | Mobility | 0.409 | 553.723 | 8.118 | 14.190 | Km | Retained |
| | Mobility index | 17.156 | 947.757 | 79.520 | 40.596 | Ratio | Retained |

| | | | | | | | |
|---|---|---|---|---|---|---|---|
| | Commute to work by public transportation | 0.000 | 61.760 | 0.948 | 3.102 | Ratio | Removed |
| | Commute to work by private vehicles | 7.840 | 99.290 | 89.715 | 6.532 | Ratio | Removed |
| | Commute to work by walking or bicycle | 0.000 | 42.410 | 3.427 | 3.146 | Ratio | Retained |
| | Leisure time physical inactivity | 9.400 | 49.800 | 26.238 | 5.498 | Ratio | Retained |
| **Environmental factors** | Developed intensity | 0.000 | 0.849 | 0.045 | 0.093 | Ratio | Removed |
| | Severe housing problems | 2.700 | 39.100 | 14.308 | 4.338 | Ratio | Retained |
| | Crowded housing | 0.000 | 16.900 | 2.314 | 1.816 | Ratio | Retained |
| | Proximity to highway | 0.000 | 16.400 | 1.943 | 1.871 | Ratio | Retained |
| | Point of interest visit (POI) | 0.000 | 10.507 | 1.505 | 0.904 | Per person | Retained |
| | Airport density | 0.000 | 0.039 | 0.002 | 0.002 | Number per $km^2$ | Retained |
| | Railway density | 0.000 | 2.625 | 0.062 | 0.111 | Length of km per $km^2$ | Retained |
| | Highway and secondary road density | 0.000 | 1.577 | 0.105 | 0.132 | Length of km per $km^2$ | Retained |
| | Sun exposure | 2856.000 | 5960.000 | 4490.738 | 452.631 | Degrees Fahrenheit | Removed |
| | Precipitation | 0.205 | 8.047 | 3.483 | 1.647 | Inch | Removed |
| | Maximum temperature | 49.682 | 91.482 | 68.836 | 7.973 | Degrees Fahrenheit | Removed |
| | Minimum temperature | 22.782 | 71.318 | 46.529 | 8.232 | Degrees Fahrenheit | Removed |
| | Relative humidity | 0.380 | 0.870 | 0.765 | 0.080 | Percent | Removed |
| | PM2.5 | 1.500 | 16.000 | 7.640 | 1.674 | Kj per $m^2$ | Retained |

| | | | | | | | |
|---|---|---|---|---|---|---|---|
| | PM10 | 7.476 | 57.922 | 17.417 | 4.755 | $Ug/m^3$ | Retained |
| | NO2 | 2.896 | 27.402 | 13.321 | 3.337 | Ppb | Retained |
| | Average temperature | 36.809 | 79.218 | 57.687 | 7.941 | Degrees Fahrenheit | Retained |
| | Wind speed | 3.969 | 9.923 | 7.026 | 0.800 | M/s | Retained |
| **Green space factors** | Developed open space | 0.001 | 0.337 | 0.045 | 0.039 | Developed open space area/ county area | Retained |
| | Forest | 0.000 | 0.929 | 0.306 | 0.256 | Forest area/county area | Retained |
| | Forest U1 | 0.000 | 0.858 | 0.303 | 0.211 | Ratio | Retained |
| | Forest U2 | 0.000 | 0.876 | 0.332 | 0.239 | Ratio | Retained |
| | Forest U3 | 0.000 | 0.906 | 0.305 | 0.239 | Ratio | Retained |
| | Forest U4 | 0.000 | 0.887 | 0.312 | 0.255 | Ratio | Retained |
| | Forest U5 | 0.000 | 0.929 | 0.296 | 0.277 | Ratio | Retained |
| | Shrub / Scrub | 0.000 | 0.976 | 0.085 | 0.181 | Shrub and scrub area/county area | Retained |
| | Grassland/ Herbaceous | 0.000 | 0.977 | 0.094 | 0.169 | Grassland and herbaceous area/county area | Retained |
| | Hay / Pasture | 0.000 | 0.799 | 0.101 | 0.123 | Pasture and hay area/county area | Retained |
| | Cultivated crops | 0.000 | 0.928 | 0.222 | 0.265 | Cultivated crops area/county area | Removed |
| | Local parks | 0.000 | 1.141 | 0.002 | 0.009 | Local park area/county area | Retained |
| | Tree canopy | 0.047 | 87.315 | 35.314 | 25.138 | Range 0-100 | Removed |
| | NDVI | 0.071 | 0.816 | 0.525 | 0.146 | Range 0-1 | Removed |

Note: Min = minimum; Max = maximum; SD = standard deviation; Per 100k = 100,000 population; USD = United States Dollar; all data are at the county level; Infection U1-5=infection rate for urbanicity level 1-5; Infection P1-5=infection rate for period 1-5; Forest U1-5=forest ratio for urbanicity level 1-5.

Table S4. **Regression results of association between green spaces and infection rate by 5 urbanicity level.**

| **Model factors** | | **Urbanicity level 1** | **Urbanicity level 2** | **Urbanicity level 3** | **Urbanicity level 4** | **Urbanicity level 5** |
|---|---|---|---|---|---|---|
| **Socioeconomic and demographic factors** | Population density | -0.007 (-0.044, 0.030) | N/A | N/A | N/A | 0.419 (-0.587, 1.425) |
| | Black non-Hispanic | -0.155 (-0.248, -0.062)* | -0.068 (-0.161, 0.026) | -0.117 (-0.253, 0.018) | 0.004 (-0.079, 0.087) | -0.016 (-0.077, 0.046) |
| | Population aged 65+ | -0.162 (-0.263, -0.061)* | -0.205 (-0.310, -0.100) ** | -0.153 (-0.247, -0.059)* | -0.201 (-0.285, -0.117) *** | -0.161 (-0.222, -0.100) *** |
| | Gini Index | 0.059 (-0.023, 0.142) | 0.168 (0.068, 0.268)* | 0.056 (-0.067, 0.179) | 0.042 (-0.033, 0.117) | 0.054 (0.004, 0.104)† |
| | Median home value | N/A | -0.195 (-0.317, -0.072)* | 0.005 (-0.188, 0.198) | 0.070 (-0.036, 0.175) | -0.015 (-0.117, 0.088) |
| | Unemployment rate | 0.014 (-0.124, 0.153) | -0.162 (-0.267, -0.057)* | 0.102 (0.001, 0.202)† | -0.043 (-0.129, 0.042) | -0.053 (-0.108, 0.002) |
| | Population without high school diploma | N/A | N/A | 0.102 (-0.063, 0.267) | 0.129 (0.035, 0.224)* | 0.061 (-0.015, 0.136) |
| **Healthcare and testing** | Population without insurance | N/A | N/A | N/A | -0.100 (-0.201, 0.001) | 0.006 (-0.064, 0.077) |
| | Testing rate | 0.320 (0.248, 0.392) *** | 0.253 (0.169, 0.336) *** | 0.163 (0.095, 0.231) *** | 0.328 (0.261, 0.396) *** | 0.437 (0.385, 0.488) *** |
| **Pre-existing chronic disease factors** | Diabetes rate | 0.000 (-0.116, 0.115) | 0.035 (-0.058, 0.128) | -0.025 (-0.140, 0.091) | 0.051 (-0.030, 0.132) | 0.004 (-0.046, 0.055) |
| | Obesity rate | 0.012 (-0.082, 0.105) | 0.027 (-0.069, 0.122) | -0.102 (-0.226, 0.021) | -0.046 (-0.128, 0.035) | -0.009 (-0.063, 0.044) |
| | Stroke mortality | 0.085 (-0.003, 0.172) | 0.009 (-0.080, 0.097) | -0.071 (-0.178, 0.035) | -0.006 (-0.076, 0.063) | 0.000 (-0.060, 0.060) |

| | | | | | | |
|---|---|---|---|---|---|---|
| | Hypertension mortality | 0.048 (-0.047, 0.143) | 0.053 (-0.022, 0.128) | -0.054 (-0.159, 0.051) | -0.007 (-0.073, 0.058) | 0.016 (-0.036, 0.068) |
| | Heart disease mortality | 0.010 (-0.089, 0.109) | -0.005 (-0.092, 0.082) | 0.079 (-0.027, 0.185) | 0.028 (-0.040, 0.095) | 0.049 (-0.007, 0.105) |
| **Environmental factors** | Severe housing problem | N/A | N/A | -0.076 (-0.216, 0.065) | -0.204 (-0.300, -0.108)*** | -0.102 (-0.168, -0.035)* |
| | Overcrowded Housing | 0.104 (0.017, 0.191)† | 0.073 (-0.028, 0.174) | 0.043 (-0.109, 0.195) | 0.147 (0.058, 0.236)* | 0.098 (0.037, 0.158)* |
| | Proximity to highway | -0.032 (-0.102, 0.039) | 0.019 (-0.065, 0.103) | 0.062 (-0.023, 0.146) | 0.079 (0.018, 0.141)† | -0.003 (-0.059, 0.054) |
| | Airport density | -0.043 (-0.081, -0.005)† | -0.025 (-0.093, 0.042) | -0.132 (-0.225, -0.038)* | -0.095 (-0.183, -0.007)† | -0.115 (-0.189, -0.040)* |
| | Railway density | -0.017 (-0.050, 0.017) | -0.100 (-0.201, 0.000) | -0.054 (-0.204, 0.096) | 0.064 (-0.093, 0.221) | -0.002 (-0.135, 0.131) |
| | Highway and secondary road density | N/A | N/A | N/A | -0.213 (-0.377, -0.049)† | 0.012 (-0.124, 0.149) |
| | PM 2.5 | 0.165 (0.088, 0.241)*** | 0.128 (0.047, 0.209)* | 0.108 (-0.012, 0.227) | 0.081 (-0.013, 0.175) | 0.017 (-0.065, 0.098) |
| | PM 10 | 0.040 (-0.040, 0.121) | N/A | -0.004 (-0.113, 0.106) | 0.214 (0.118, 0.309)*** | 0.198 (0.118, 0.277)*** |
| | Average temperature | -0.136 (-0.244, -0.028)† | -0.196 (-0.306, -0.087)** | N/A | N/A | N/A |
| | Wind speed | 0.037 (-0.052, 0.126) | -0.019 (-0.096, 0.058) | 0.058 (-0.044, 0.159) | 0.077 (0.004, 0.150)† | 0.027 (-0.032, 0.086) |
| | $NO_2$ | 0.074 (-0.001, 0.149) | 0.008 (-0.082, 0.099) | -0.074 (-0.177, 0.029) | -0.046 (-0.122, 0.030) | -0.043 (-0.109, 0.023) |
| **Behavioral factor** | Commute to work by walking or bicycle | -0.243 (-0.349, -0.137)*** | -0.149 (-0.277, -0.022)† | -0.149 (-0.265, -0.034)† | -0.127 (-0.211, -0.043)* | -0.061 (-0.109, -0.014)† |
| | Leisure time physical inactivity | 0.004 (-0.100, 0.108) | 0.002 (-0.109, 0.112) | -0.036 (-0.157, 0.085) | -0.018 (-0.106, 0.069) | -0.002 (-0.061, 0.058) |

| | | | | | | |
|---|---|---|---|---|---|---|
| | Smoker | 0.197 (0.093, 0.300)** | -0.003 (-0.113, 0.107) | 0.069 (-0.058, 0.195) | 0.093 (-0.005, 0.192) | 0.124 (0.055, 0.194)** |
| | Essential worker | N/A | 0.059 (-0.075, 0.192) | 0.270 (0.108, 0.431)* | 0.073 (-0.030, 0.175) | 0.010 (-0.059, 0.079) |
| | POI visits | 0.028 (-0.035, 0.091) | 0.003 (-0.085, 0.091) | 0.089 (-0.001, 0.179) | 0.088 (0.020, 0.157)† | 0.067 (0.018, 0.117)* |
| | Mobility | 0.280 (-0.029, 0.589) | 0.089 (-0.046, 0.224) | -0.106 (-0.463, 0.252) | 0.138 (-0.183, 0.458) | -0.002 (-0.033, 0.029) |
| | Mobility 50 index | -0.005 (-0.114, 0.103) | 0.291 (0.030, 0.551)† | 0.377 (0.102, 0.652)* | 0.288 (0.179, 0.397)*** | -0.003 (-0.037, 0.031) |
| **Policy and regulation** | State governor party | -0.022 (-0.102, 0.058) | 0.009 (-0.087, 0.105) | -0.083 (-0.208, 0.041) | 0.041 (-0.046, 0.127) | 0.069 (-0.008, 0.145) |
| | Stay-at-home orders | 0.031 (-0.030, 0.092) | 0.039 (-0.029, 0.108) | -0.059 (-0.164, 0.045) | 0.036 (-0.028, 0.099) | 0.013 (-0.046, 0.072) |
| | Public mask mandates | -0.051 (-0.141, 0.039) | -0.136 (-0.237, -0.035)* | -0.081 (-0.208, 0.046) | -0.232 (-0.322, -0.142) *** | -0.190 (-0.264, -0.116) *** |
| | Business closing and reopening | -0.075 (-0.154, 0.004) | -0.089 (-0.174, -0.005)† | -0.224 (-0.328, -0.120) *** | -0.113 (-0.184, -0.041)* | -0.151 (-0.204, -0.099) *** |
| | Developed open space | -0.018 (-0.067, 0.031) | -0.015 (-0.107, 0.076) | 0.014 (-0.155, 0.183) | 0.229 (0.060, 0.397)* | -0.270 (-0.339, -0.200) *** |
| **Green space factors** | Forest | -0.209 (-0.299, -0.118) *** | -0.274 (-0.355, -0.194) *** | -0.431 (-0.550, -0.311) *** | -0.267 (-0.345, -0.188) *** | -0.108 (-0.169, -0.047)** |
| | Shrub / Scrub | -0.103 (-0.223, 0.018) | -0.023 (-0.129, 0.082) | -0.082 (-0.191, 0.027) | -0.031 (-0.112, 0.050) | 0.002 (-0.051, 0.055) |
| | Grassland / Herbaceous | 0.005 (-0.102, 0.113) | -0.036 (-0.144, 0.073) | -0.029 (-0.138, 0.080) | -0.012 (-0.094, 0.071) | -0.087 (-0.139, -0.034)* |
| | Hay / Pasture | -0.040 (-0.110, 0.029) | -0.103 (-0.175, -0.032)* | -0.129 (-0.223, -0.034)* | -0.067 (-0.133, 0.000)† | -0.110 (-0.220, -0.001)† |
| | Local parks | 0.007 (-0.032, 0.047) | -0.028 (-0.163, 0.108) | -0.023 (-0.206, 0.159) | 0.071 (-0.137, 0.279) | -0.270 (-0.339, -0.200) |

| | | | | | | |
|---|---|---|---|---|---|---|
| | | | | | | *** |
| | Adjusted $R^2$ without green space factors | 0.409, <0.0001*** | 0.354, <0.0001*** | 0.402, <0.0001*** | 0.527, <0.0001*** | 0.463, <0.0001*** |
| | Adjusted $R^2$ | 0.438, <0.0001*** | 0.444, <0.0001*** | 0.494, <0.0001*** | 0.564, <0.0001*** | 0.496, <0.0001*** |
| | $R^2$change and ANOVA F-statistic | 0.030, 0.0002** | 0.090, 10.096*** | 0.092, 10.771*** | 0.037, 9.564*** | 0.033,17.642*** |

Note: † indicates $p$< 0.05; * indicates $p$ < 0.01; ** indicates $p$ < 0.001; *** indicates $p$ < 0.0001; each cell indicates β value, 95% CI, $p$ value.

Table S5. **Regression results of association between green spaces and infection rate by five time periods.**

| **Model factors** | | **Period 1** | **Period 2** | **Period 3** | **Period 4** | **Period 5** |
|---|---|---|---|---|---|---|
| **Socioeconomic and demographic factors** | Population density | 0.201 (0.157, 0.245) *** | 0.004 (-0.038, 0.046) | -0.034 (-0.068, 0.001) | 0.005 (-0.037, 0.048) | 0.057 (0.020, 0.094)* |
| | Black non-Hispanic | 0.149 (0.104, 0.194) *** | 0.169 (0.126, 0.212) *** | 0.252 (0.218, 0.287) *** | 0.050 (0.007, 0.093)† | -0.157 (-0.194, -0.119) *** |
| | Population aged 65+ | -0.014 (-0.052, 0.024) | -0.056 (-0.092, -0.019)* | -0.057 (-0.087, -0.027) ** | -0.091 (-0.128, -0.053) *** | -0.082 (-0.115, -0.050) *** |
| | Gini Index | 0.048 (0.011, 0.085)† | -0.053 (-0.089, -0.017)* | 0.015 (-0.015, 0.045) | 0.086 (0.050, 0.122) *** | 0.061 (0.029, 0.092)** |
| | Median home value | 0.209 (0.157, 0.261) *** | 0.031 (-0.020, 0.082) | 0.017 (-0.025, 0.058) | -0.025 (-0.076, 0.025) | -0.068 (-0.113, -0.024)* |
| | Unemployment rate | 0.002 (-0.039, 0.042) | -0.026 (-0.065, 0.013) | 0.040 (0.007, 0.072)† | -0.091 (-0.130, -0.052) *** | -0.030 (-0.064, 0.004) |
| | Population without high school diploma | -0.005 (-0.060, 0.051) | 0.158 (0.105, 0.212) *** | 0.153 (0.109, 0.196) *** | 0.095 (0.043, 0.148)** | 0.020 (-0.027, 0.066) |
| **Healthcare and testing** | Population without insurance | 0.020 (-0.033, 0.072) | 0.139 (0.087, 0.191) *** | 0.119 (0.079, 0.160) *** | 0.091 (0.041, 0.141)** | 0.024 (-0.021, 0.069) |
| | Testing rate | 0.310 (0.280, 0.340) *** | 0.472 (0.442, 0.503) *** | 0.291 (0.266, 0.316) *** | 0.268 (0.236, 0.299) *** | 0.285 (0.256, 0.314) *** |
| **Pre-existing chronic disease factors** | Diabetes rate | -0.009 (-0.048, 0.030) | -0.017 (-0.055, 0.020) | -0.023 (-0.054, 0.008) | 0.009 (-0.028, 0.047) | 0.029 (-0.004, 0.063) |
| | Obesity rate | -0.067 (-0.107, -0.026)* | -0.051 (-0.090, -0.013)* | -0.002 (-0.033, 0.030) | -0.044 (-0.083, -0.005)† | 0.003 (-0.031, 0.037) |
| | Stroke mortality | -0.075 (-0.115, -0.035) ** | -0.039 (-0.077, 0.000)† | 0.032 (0.001, 0.064)† | 0.048 (0.009, 0.086)† | 0.009 (-0.024, 0.043) |
| | Hypertension mortality | 0.042 | 0.020 | -0.014 | -0.018 | 0.026 |

| | | | | | | |
|---|---|---|---|---|---|---|
| | | (0.006, 0.078)† | (-0.015, 0.055) | (-0.043, 0.014) | (-0.053, 0.017) | (-0.005, 0.057) |
| | Heart disease mortality | -0.051<br>(-0.089, -0.013)* | -0.039<br>(-0.076, -0.002)† | 0.004<br>(-0.027, 0.034) | 0.047<br>(0.010, 0.084)† | 0.019<br>(-0.013, 0.052) |
| **Environmental factors** | Severe housing problem | 0.022<br>(-0.024, 0.068) | 0.032<br>(-0.013, 0.076) | 0.007<br>(-0.030, 0.044) | -0.121<br>(-0.166, -0.077)<br>*** | -0.145<br>(-0.184, -0.106)<br>*** |
| | Overcrowded Housing | 0.060<br>(0.014, 0.106)† | 0.048<br>(0.004, 0.092)† | 0.168<br>(0.131, 0.204)<br>*** | 0.074<br>(0.031, 0.118)** | 0.012<br>(-0.027, 0.050) |
| | Proximity to highway | -0.002<br>(-0.037, 0.033) | 0.049<br>( 0.015, 0.082)* | -0.014<br>(-0.042, 0.014) | -0.018<br>(-0.051, 0.016) | 0.013<br>(-0.017, 0.042) |
| | Airport density | -0.081<br>(-0.114, -0.048)<br>*** | -0.023<br>(-0.055, 0.008) | -0.022<br>(-0.049, 0.004) | -0.073<br>(-0.105, -0.041)<br>*** | -0.059<br>(-0.087, -0.031)<br>*** |
| | Railway density | -0.008<br>(-0.048, 0.032) | -0.005<br>(-0.044, 0.033) | -0.042<br>(-0.074, -0.010)* | -0.057<br>(-0.096, -0.019)* | -0.015<br>(-0.049, 0.019) |
| | Highway and secondary road density | -0.014<br>(-0.068, 0.040) | -0.022<br>(-0.074, 0.030) | -0.025<br>(-0.068, 0.018) | -0.029<br>(-0.081, 0.023) | -0.048<br>(-0.093, -0.002)† |
| | PM 2.5 | -0.021<br>(-0.067, 0.025) | 0.048<br>(0.003, 0.092)† | 0.065<br>(0.028, 0.102)** | 0.026<br>(-0.020, 0.071) | 0.096<br>(0.056, 0.136)<br>*** |
| | PM 10 | -0.022<br>(-0.062, 0.018) | -0.021<br>(-0.060, 0.019) | 0.079<br>(0.042, 0.115)<br>*** | 0.070<br>(0.026, 0.113)* | 0.169<br>(0.135, 0.202)<br>*** |
| | Average temperature | -0.088<br>(-0.144, -0.032)* | -0.124<br>(-0.177, -0.072)<br>*** | 0.075<br>(0.032, 0.117)** | -0.179<br>(-0.232, -0.127)<br>*** | -0.346<br>(-0.389, -0.302)<br>*** |
| | Wind speed | 0.087<br>(0.049, 0.126)*** | 0.045<br>(0.008, 0.083)† | -0.105<br>(-0.136, -0.074)<br>*** | 0.015<br>(-0.022, 0.053) | 0.030<br>(-0.002, 0.062) |
| | $NO_2$ | 0.136<br>(0.096, 0.177)*** | 0.093<br>(0.055, 0.131)*** | -0.011<br>(-0.041, 0.018) | -0.043<br>(-0.080, -0.005)† | -0.014<br>(-0.047, 0.020) |
| **Behavior factor** | Commute to work by walking or bicycle | -0.021<br>(-0.057, 0.016) | -0.023<br>(-0.059, 0.012) | -0.027<br>(-0.056, 0.003) | 0.020<br>(-0.016, 0.055) | -0.098<br>(-0.129, -0.067)<br>*** |
| | Leisure time physical inactivity | 0.079<br>(0.035, 0.123)** | 0.054<br>(0.012, 0.097)† | 0.027<br>(-0.007, 0.062) | 0.037<br>(-0.005, 0.080) | 0.022<br>(-0.015, 0.059) |
| | Smoker | -0.040<br>(-0.085, 0.006) | -0.065<br>(-0.110, -0.020)* | -0.040<br>(-0.079, -0.001) | 0.113<br>(0.067, 0.160) | 0.089<br>(0.048, 0.130) |

| | | | | | | |
|---|---|---|---|---|---|---|
| | | | | † | *** | *** |
| | Essential worker | 0.062 (0.013, 0.111)† | 0.051 (0.005, 0.098)† | -0.051 (-0.090, -0.013)* | 0.020 (-0.027, 0.067) | 0.086 (0.045, 0.127)*** |
| | POI visits | 0.011 (-0.022, 0.045) | -0.058 (-0.090, -0.025)** | 0.056 (0.029, 0.082) *** | 0.070 (0.038, 0.102) *** | 0.107 (0.079, 0.135) *** |
| | Mobility | 0.003 (-0.027, 0.033) | -0.015 (-0.044, 0.014) | -0.017 (-0.041, 0.007) | -0.009 (-0.038, 0.020) | 0.001 (-0.025, 0.026) |
| | Mobility 50 index | -0.006 (-0.035, 0.024) | 0.011 (-0.022, 0.044) | 0.007 (-0.019, 0.032) | -0.028 (-0.058, 0.003) | 0.010 (-0.016, 0.037) |
| **Policy and regulation** | State governor party | -0.018 (-0.056, 0.021) | 0.021 (-0.019, 0.060) | -0.001 (-0.039, 0.037) | 0.041 (0.000, 0.083) | -0.036 (-0.070, -0.001)† |
| | Stay-at-home orders | -0.040 (-0.079, -0.001)† | 0.001 (-0.040, 0.041) | 0.001 (-0.025, 0.028) | 0.086 (0.055, 0.117) *** | 0.125 (0.097, 0.152) *** |
| | Public mask mandates | N/A | 0.016 (-0.020, 0.053) | -0.178 (-0.216, -0.139) *** | -0.233 (-0.273, -0.192) *** | -0.001 (-0.035, 0.033) |
| | Business closing and reopening | -0.003 (-0.042, 0.036) | 0.058 (0.025, 0.092)** | 0.039 (0.012, 0.066)* | -0.150 (-0.185, -0.115) *** | -0.047 (-0.077, -0.017)* |
| | Developed open space | 0.008 (-0.042, 0.058) | 0.034 (-0.014, 0.082) | 0.019 (-0.021, 0.059) | 0.024 (-0.024, 0.072) | 0.077 (0.035, 0.120)** |
| **Green space factors** | Forest | -0.021 (-0.061, 0.018) | -0.102 (-0.141, -0.063) *** | -0.066 (-0.100, -0.033) ** | -0.162 (-0.202, -0.122) *** | -0.243 (-0.278, -0.208) *** |
| | Shrub / Scrub | -0.085 (-0.127, -0.043)** | -0.106 (-0.145, -0.066) *** | -0.035 (-0.068, -0.002)† | -0.072 (-0.114, -0.031) ** | 0.007 (-0.029, 0.043) |
| | Grassland / Herbaceous | -0.034 (-0.074, 0.007) | -0.024 (-0.062, 0.013) | -0.059 (-0.090, -0.027) ** | 0.001 (-0.038, 0.041) | 0.128 (0.094, 0.161) *** |
| | Hay / Pasture | 0.010 (-0.025, 0.045) | -0.030 (-0.064, 0.004) | -0.024 (-0.051, 0.004) | -0.028 (-0.062, 0.005) | -0.039 (-0.068, -0.010)* |
| | Local parks | 0.010 (-0.033, 0.054) | 0.031 (-0.010, 0.072) | -0.064 (-0.098, -0.030) ** | -0.037 (-0.079, 0.004) | -0.046 (-0.082, -0.009)† |
| | Adjusted $R^2$ without green | 0.342, <0.0001*** | 0.384, <0.0001*** | 0.585, | 0.375, | 0.482, <0.0001 |

| | | | | | | |
|---|---|---|---|---|---|---|
| | space factors | | | <0.0001*** | <0.0001*** | |
| | Adjusted $\boldsymbol{R^2}$ | 0.345, <0.0001*** | 0.394, <0.0001*** | 0.590, <0.0001*** | 0.390, <0.0001*** | 0.532, <0.0001*** |
| | $\boldsymbol{R^2}$ change and ANOVA F-statistic | 0.004, 3.720*** | 0.011, 9.973*** | 0.005, 6.908*** | 0.015, 13.783*** | 0.050, 55.595*** |

Note: † indicates $p < 0.05$; * indicates $p < 0.01$; ** indicates $p < 0.001$; *** indicates $p < 0.0001$; each cell indicates β value, 95% CI, *p* value.

Table S6. **Regression results of association between the ratio of forest and infection rate across five periods and five urbanicity levels.**

| **Forest** | **Period 1** | **Period 2** | **Period 3** | **Period 4** | **Period 5** |
|---|---|---|---|---|---|
| Urbanicity level 1 | -0.010 (-0.232, 0.211) | -0.123 (-0.283, 0.038) | 0.023 (-0.044, 0.089) | -0.061 (-0.116, -0.006)† | -0.179 (-0.249, -0.109) *** |
| Urbanicity level 2 | -0.028 (-0.087, 0.030) | -0.056 (-0.120, 0.008) | -0.029 (-0.119, 0.061) | -0.028 (-0.123, 0.066) | -0.270 (-0.338, -0.201) *** |
| Urbanicity level 3 | 0.005 (-0.069, 0.079) | -0.036 (-0.182, 0.110) | -0.085 (-0.172, 0.002) | -0.256 (-0.380, -0.132) ** | -0.244 (-0.349, -0.140) *** |
| Urbanicity level 4 | -0.028 (-0.105, 0.050) | -0.041 (-0.142, 0.060) | -0.077 (-0.148, -0.007)† | -0.144 (-0.224, -0.065) ** | -0.216 (-0.287, -0.145) *** |
| Urbanicity level 5 | 0.052 (0.013, 0.090)* | -0.069 (-0.122, -0.015)† | -0.101 (-0.163, -0.039)* | -0.157 (-0.233, -0.081) ** | -0.199 (-0.268, -0.129) *** |

Note: † indicates $p < 0.05$; * indicates $p < 0.01$; ** indicates $p < 0.001$; *** indicates $p < 0.0001$; each cell indicates β value, 95% CI, *p* value.

Table S7. **Regression results of association between population-weighted forest exposure and infection rate of varying buffer size.**

| **Buffer distance of forest** | **Beta value by forest** | **Beta value difference by forest** | **Adj. $R^2$ change by forest** | **$R^2$ without forest exposure** | **$R^2$ change** | **Adj. R2 change difference by forest** |
|---|---|---|---|---|---|---|
| 100m | -0.2500 | -0.0093 | 0.4930 | 0.4645 | 0.0285 | 0.0018 |
| 200m | -0.2593 | -0.0042 | 0.4948 | 0.4645 | 0.0303 | 0.0009 |
| 300m | -0.2635 | -0.0027 | 0.4957 | 0.4645 | 0.0312 | 0.0005 |
| 400m | -0.2662 | -0.0024 | 0.4962 | 0.4645 | 0.0317 | 0.0006 |
| 500m | -0.2686 | -0.0012 | 0.4968 | 0.4645 | 0.0323 | 0.0002 |
| 600m | -0.2698 | -0.0009 | 0.4970 | 0.4645 | 0.0325 | 0.0002 |
| 700m | -0.2707 | -0.0008 | 0.4972 | 0.4645 | 0.0327 | 0.0002 |
| 800m | -0.2715 | -0.0006 | 0.4974 | 0.4645 | 0.0329 | 0.0002 |
| 900m | -0.2721 | -0.0004 | 0.4976 | 0.4645 | 0.0331 | 0.0001 |
| 1.0km | -0.2725 | -0.0002 | 0.4977 | 0.4645 | 0.0332 | 0.0001 |
| 1.1km | -0.2727 | -0.0002 | 0.4978 | 0.4645 | 0.0333 | 0.0000 |
| 1.2km | -0.2729 | -0.0001 | 0.4978 | 0.4645 | 0.0333 | 0.0001 |
| 1.3km | -0.2730 | 0.0000 | 0.4979 | 0.4645 | 0.0334 | 0.0000 |
| 1.4km | -0.2730 | 0.0001 | 0.4979 | 0.4645 | 0.0334 | 0.0001 |
| 1.5km | -0.2729 | 0.0001 | 0.4980 | 0.4645 | 0.0335 | 0.0000 |
| 1.6km | -0.2728 | 0.0001 | 0.4980 | 0.4645 | 0.0335 | 0.0000 |
| 1.7km | -0.2727 | 0.0001 | 0.4980 | 0.4645 | 0.0335 | 0.0000 |
| 1.8km | -0.2726 | 0.0002 | 0.4980 | 0.4645 | 0.0335 | 0.0000 |
| 1.9km | -0.2724 | 0.0002 | 0.4980 | 0.4645 | 0.0335 | 0.0000 |
| 2.0km | -0.2722 | 0.0001 | 0.4980 | 0.4645 | 0.0335 | 0.0000 |
| 2.1km | -0.2721 | 0.0002 | 0.4980 | 0.4645 | 0.0335 | 0.0000 |
| 2.2km | -0.2719 | 0.0001 | 0.4980 | 0.4645 | 0.0335 | 0.0000 |
| 2.3km | -0.2718 | 0.0002 | 0.4980 | 0.4645 | 0.0335 | 0.0000 |
| 2.4km | -0.2716 | 0.0001 | 0.4980 | 0.4645 | 0.0335 | 0.0000 |
| 2.5km | -0.2715 | 0.0001 | 0.4980 | 0.4645 | 0.0335 | 0.0000 |
| 2.6km | -0.2714 | 0.0001 | 0.4980 | 0.4645 | 0.0335 | 0.0000 |
| 2.7km | -0.2713 | 0.0001 | 0.4980 | 0.4645 | 0.0335 | 0.0000 |
| 2.8km | -0.2712 | 0.0001 | 0.4980 | 0.4645 | 0.0335 | 0.0000 |
| 2.9km | -0.2711 | 0.0001 | 0.4980 | 0.4645 | 0.0335 | 0.0000 |
| 3.0km | -0.2710 | 0.0000 | 0.4980 | 0.4645 | 0.0335 | 0.0000 |
| 3.1km | -0.2710 | 0.0001 | 0.4980 | 0.4645 | 0.0335 | 0.0000 |
| 3.2km | -0.2709 | 0.0000 | 0.4980 | 0.4645 | 0.0335 | 0.0000 |
| 3.3km | -0.2709 | 0.0001 | 0.4980 | 0.4645 | 0.0335 | 0.0000 |
| 3.4km | -0.2708 | 0.0001 | 0.4980 | 0.4645 | 0.0335 | 0.0000 |
| 3.5km | -0.2707 | 0.0000 | 0.4980 | 0.4645 | 0.0335 | 0.0000 |
| 3.6km | -0.2707 | 0.0001 | 0.4980 | 0.4645 | 0.0335 | 0.0001 |
| 3.7km | -0.2706 | 0.0001 | 0.4981 | 0.4645 | 0.0336 | 0.0000 |
| 3.8km | -0.2710 | 0.0001 | 0.4980 | 0.4650 | 0.0340 | 0.0000 |
| 3.9km | -0.2700 | 0.0001 | 0.4980 | 0.4650 | 0.0340 | 0.0000 |
| 4.0km | -0.2700 | 0.0000 | 0.4980 | 0.4650 | 0.0340 | 0.0000 |
| 4.1km | -0.2700 | 0.0001 | 0.4980 | 0.4650 | 0.0340 | 0.0000 |
| 4.2km | -0.2702 | 0.0001 | 0.4981 | 0.4645 | 0.0336 | 0.0000 |
| 4.3km | -0.2701 | 0.0000 | 0.4981 | 0.4645 | 0.0336 | 0.0000 |
| 4.4km | -0.2701 | 0.0001 | 0.4981 | 0.4645 | 0.0336 | 0.0000 |
| 4.5km | -0.2700 | 0.0000 | 0.4981 | 0.4645 | 0.0336 | 0.0000 |
| 4.6km | -0.2700 | 0.0001 | 0.4981 | 0.4645 | 0.0336 | 0.0000 |
| 4.7km | -0.2699 | 0.0000 | 0.4981 | 0.4645 | 0.0336 | 0.0000 |
| 4.8km | -0.2699 | 0.0000 | 0.4981 | 0.4645 | 0.0336 | 0.0000 |

| | | | | | | |
|---|---|---|---|---|---|---|
| 4.9km | -0.2699 | 0.0000 | 0.4981 | 0.4645 | 0.0336 | 0.0000 |
| 5km | -0.2699 | 0.0008 | 0.4981 | 0.4645 | 0.0336 | 0.0000 |
| 6km | -0.2691 | 0.0000 | 0.4981 | 0.4645 | 0.0336 | 0.0001 |
| 7km | -0.2691 | -0.0006 | 0.4982 | 0.4645 | 0.0337 | 0.0002 |
| 8km | -0.2697 | -0.0007 | 0.4984 | 0.4645 | 0.0339 | 0.0002 |
| 9km | -0.2704 | -0.0009 | 0.4986 | 0.4645 | 0.0341 | 0.0002 |
| 10km | -0.2713 | -0.0042 | 0.4988 | 0.4645 | 0.0343 | 0.0010 |
| 15km | -0.2755 | -0.0026 | 0.4998 | 0.4645 | 0.0353 | 0.0004 |
| 20km | -0.2781 | -0.0022 | 0.5002 | 0.4645 | 0.0357 | 0.0002 |
| 25km | -0.2803 | -0.0015 | 0.5004 | 0.4645 | 0.0359 | 0.0000 |
| 30km | -0.2818 | -0.0015 | 0.5004 | 0.4645 | 0.0359 | -0.0001 |
| 35km | -0.2833 | 0.5685 | 0.5003 | 0.4645 | 0.0358 | 0.0001 |
| 40km | 0.2852 | -0.5729 | 0.5004 | 0.4645 | 0.0359 | 0.0002 |
| 45km | -0.2877 | -0.0026 | 0.5006 | 0.4645 | 0.0361 | 0.0002 |
| 50km | -0.2903 | 0.2903 | 0.5008 | 0.4645 | 0.0363 | 0.0002 |

Note: All $p$ values are <0.0001 in this table.

Table S8. **Mean forest exposure per capita and population-weighted forest exposure by five urbanicity levels.**

| **Urbanicity level** | **Mean forest exposure per capita ($km^2$)** | **Standard deviation of forest exposure per capita ($km^2$)** |
|---|---|---|
| Urbanicity level 1 | 1032.598 | 3673.324 |
| Urbanicity level 2 | 1356.383 | 3948.200 |
| Urbanicity level 3 | 2111.222 | 5588.354 |
| Urbanicity level 4 | 2885.554 | 8651.686 |
| Urbanicity level 5 | 8969.556 | 23106.289 |

Table S9. **Descriptive data for confounding variables at five urbanicity levels.**

| **Urbanicity level** | **Urbanicity level 1** | **Urbanicity level 2** | **Urbanicity level 3** | **Urbanicity level 4** | **Urbanicity level 5** |
|---|---|---|---|---|---|
| Population density | 485.038 | 105.780 | 65.423 | 26.298 | 11.277 |
| Female household ratio | 11.746 | 12.118 | 11.482 | 11.528 | 10.241 |
| White non-Hispanic | 70.689 | 73.636 | 77.480 | 76.948 | 79.445 |
| Black or African American non-Hispanic | 12.038 | 10.937 | 9.330 | 8.143 | 7.669 |
| Hispanic/Latino | 10.767 | 10.360 | 8.348 | 10.369 | 8.132 |
| Population aged 65 and older | 6626.100 | 6223.700 | 6144.500 | 11260.600 | 27018.300 |
| Median household income | 70119.266 | 56398.374 | 53371.831 | 50186.813 | 46803.204 |
| Gini Index | 0.440 | 0.448 | 0.447 | 0.448 | 0.446 |
| Median housing value | 237027.523 | 166569.106 | 153546.479 | 137370.487 | 112460.105 |
| Poverty rate | 10.893 | 13.998 | 14.815 | 15.900 | 16.687 |
| Food stamp supplement | 11.188 | 13.929 | 13.853 | 14.943 | 14.471 |
| Unemployment rate | 3.692 | 4.078 | 4.076 | 4.220 | 4.173 |
| Population without college degree | 69.743 | 75.067 | 76.395 | 79.603 | 82.300 |
| Population without a high school diploma | 10.935 | 12.481 | 12.215 | 13.938 | 14.652 |
| Health insurance coverage | 9.504 | 10.524 | 10.518 | 11.246 | 12.633 |
| Testing rate | 187.532 | 176.042 | 192.356 | 168.055 | 157.826 |
| Diagnosed diabetes | 9.790 | 10.386 | 10.776 | 10.832 | 10.545 |
| Obesity | 30.554 | 32.453 | 33.037 | 33.480 | 33.154 |
| Stroke mortality | 38.294 | 40.025 | 39.672 | 40.284 | 39.980 |
| Hypertension morality | 118.571 | 135.301 | 126.034 | 136.202 | 134.696 |
| Heart failure mortality | 93.875 | 102.086 | 105.544 | 110.537 | 114.539 |
| Presidential vote rates | 0.457 | 0.401 | 0.366 | 0.336 | 0.276 |
| State Governor Party | 0.530 | 0.461 | 0.434 | 0.421 | 0.404 |
| Stay-at-home orders | 4.600 | 4.548 | 4.463 | 4.442 | 4.352 |
| Public mask mandates | 0.603 | 0.518 | 0.502 | 0.497 | 0.454 |
| Business closing and reopening orders | 6.472 | 6.508 | 6.353 | 6.383 | 6.204 |
| Severe housing problem | 16.140 | 15.334 | 14.921 | 14.864 | 12.974 |
| Proximity to highway | 2.659 | 2.428 | 2.457 | 2.301 | 1.255 |
| Crowded housing | 2.377 | 2.420 | 2.244 | 2.456 | 2.214 |
| Point of interest visit (POI) | 1.524 | 1.589 | 1.590 | 1.689 | 1.362 |
| Density of airports | 0.004 | 0.003 | 0.002 | 0.002 | 0.001 |
| Density of railway | 0.141 | 0.082 | 0.070 | 0.053 | 0.032 |
| Density of highway and secondary road | 0.219 | 0.141 | 0.118 | 0.090 | 0.062 |
| Average sunlight exposure | 4410.351 | 4522.764 | 4455.234 | 4471.978 | 4527.188 |
| PM2. | 8.449 | 8.359 | 7.859 | 7.649 | 7.105 |
| PM10 | 16.794 | 17.196 | 17.310 | 17.378 | 17.734 |
| Precipitation | 3.954 | 3.867 | 3.614 | 3.435 | 3.207 |
| Average temperature | 59.679 | 59.960 | 58.203 | 56.964 | 56.595 |
| Maximum temperature | 70.012 | 70.775 | 69.168 | 68.106 | 68.165 |
| Minimum temperature | 49.338 | 49.139 | 47.231 | 45.815 | 45.017 |
| Relative humidity | 0.781 | 0.776 | 0.771 | 0.763 | 0.756 |
| Wind speed | 6.829 | 6.775 | 6.895 | 7.053 | 7.183 |
| Mean NO2 Concentration in each county, 2020 | 15.955 | 13.703 | 13.408 | 13.222 | 12.362 |
| Commute to work by public transportation | 3.229 | 0.909 | 0.799 | 0.595 | 0.412 |
| Commute to work by private vehicles | 88.710 | 91.320 | 90.680 | 90.926 | 88.747 |
| Commute to work by walk or bicycle | 2.415 | 2.552 | 3.260 | 3.308 | 4.112 |
| Leisure-time physical inactivity | 23.958 | 25.085 | 25.871 | 26.655 | 27.218 |

| | | | | | |
|---|---|---|---|---|---|
| Smoker | 15.935 | 16.977 | 17.418 | 17.954 | 17.841 |
| Essential worker | 0.454 | 0.498 | 0.514 | 0.546 | 0.552 |
| Mobility | 6.968 | 7.626 | 7.303 | 6.155 | 9.815 |
| Mobility index | 56.444 | 64.658 | 73.024 | 81.415 | 92.216 |

Table S10. **Regression results of association between the ratio of developed open space and infection rate across five time periods and five urbanicity levels.**

| **Developed open space** | **Period 1** | **Period 2** | **Period 3** | **Period 4** | **Period 5** |
|---|---|---|---|---|---|
| Urbanicity level 1 | -0.045 (-0.171, 0.080) | 0.067 (-0.017, 0.151) | -0.012 (-0.049, 0.024) | -0.025 (-0.057, 0.007) | -0.028 (-0.067, 0.011) |
| Urbanicity level 2 | 0.056 (-0.008, 0.121) | 0.042 (-0.028, 0.111) | -0.028 (-0.121, 0.064) | -0.037 (-0.142, 0.069) | 0.032 (-0.043, 0.108) |
| Urbanicity level 3 | 0.036 (-0.076, 0.147) | -0.074 (-0.284, 0.136) | 0.144 (0.026, 0.263)† | -0.089 (-0.269, 0.092) | 0.060 (-0.083, 0.204) |
| Urbanicity level 4 | -0.137 (-0.317, 0.044) | 0.168 (-0.046, 0.382) | 0.103 (-0.050, 0.255) | 0.191 (0.018, 0.364)† | 0.133 (-0.025, 0.290) |
| Urbanicity level 5 | N/A | N/A | N/A | N/A | N/A |

Note: † indicates $p< 0.05$; * indicates $p < 0.01$; ** indicates $p < 0.001$; *** indicates $p < 0.0001$; each cell indicates $\beta$ value, 95% CI, $p$ value.

Table S11. **Regression results of association between the ratio of shrub/scrub and infection rate across five time periods and urbanicity levels.**

| **Shrub / scrub** | **Period 1** | **Period 2** | **Period 3** | **Period 4** | **Period 5** |
|---|---|---|---|---|---|
| Urbanicity level 1 | -0.534 (-0.839,-0.229) ** | 0.030 (-0.178, 0.238) | 0.067 (-0.023, 0.157) | -0.064 (-0.143, 0.016) | -0.119 (-0.217, -0.020)† |
| Urbanicity level 2 | -0.035 (-0.109, 0.039) | 0.019 (-0.058, 0.096) | -0.096 (-0.200, 0.009) | -0.006 (-0.133, 0.121) | -0.002 (-0.093, 0.090) |
| Urbanicity level 3 | -0.090 (-0.161, -0.018)† | -0.118 (-0.255, 0.020) | 0.018 (-0.062, 0.098) | -0.167 (-0.288, -0.046)* | -0.005 (-0.100, 0.091) |
| Urbanicity level 4 | -0.041 (-0.124, 0.042) | -0.246 (-0.347, -0.145) *** | -0.020 (-0.090, 0.050) | -0.112 (-0.197, -0.027)† | 0.053 (-0.023, 0.130) |
| Urbanicity level 5 | -0.030 (-0.066, 0.006) | -0.063 (-0.112, -0.015)* | -0.056 (-0.112, 0.000) | -0.133 (-0.202, -0.063) ** | -0.050 (-0.114, 0.013) |

Note: † indicates $p < 0.05$; * indicates $p < 0.01$; ** indicates $p < 0.001$; *** indicates $p < 0.0001$; each cell indicates $\beta$ value, 95% CI, $p$ value.

Table S12. **Regression results of association between the ratio of grassland/herbaceous and infection rate across five time periods and five urbanicity levels.**

| Grassland/ herbaceous | Period 1 | Period 2 | Period 3 | Period 4 | Period 5 |
|---|---|---|---|---|---|
| Urbanicity level 1 | -0.117 (-0.384, 0.150) | 0.129 (-0.052, 0.310) | -0.062 (-0.142, 0.018) | -0.023 (-0.095, 0.049) | 0.034 (-0.053, 0.121) |
| Urbanicity level 2 | 0.102 (0.023, 0.180)† | 0.004 (-0.079, 0.087) | -0.047 (-0.159, 0.064) | -0.001 (-0.126, 0.124) | 0.026 (-0.065, 0.118) |
| Urbanicity level 3 | -0.028 (-0.103, 0.047) | -0.078 (-0.219, 0.064) | -0.014 (-0.093, 0.064) | -0.035 (-0.157, 0.087) | 0.107 (0.011, 0.203)† |
| Urbanicity level 4 | -0.090 (-0.180, 0.001) | -0.035 (-0.142, 0.071) | -0.049 (-0.123, 0.025) | -0.055 (-0.143, 0.033) | 0.088 (0.011, 0.164)† |
| Urbanicity level 5 | -0.023 (-0.056, 0.011) | -0.036 (-0.080, 0.007) | -0.062 (-0.113, -0.011)† | -0.030 (-0.094, 0.033) | 0.091 (0.036, 0.146)* |

Note: † indicates $p < 0.05$; * indicates $p < 0.01$; ** indicates $p < 0.001$; *** indicates $p < 0.0001$; each cell indicates $\beta$ value, 95% CI, $p$ value.

Table S13. **Regression results of association between the ratio of hay/pasture infection rate across five time periods and five urbanicity levels.**

| **Hay / pasture** | **Period 1** | **Period 2** | **Period 3** | **Period 4** | **Period 5** |
|---|---|---|---|---|---|
| Urbanicity level 1 | -0.037 (-0.223, 0.149) | 0.039 (-0.084, 0.162) | -0.005 (-0.057, 0.047) | -0.010 (-0.055, 0.035) | -0.045 (-0.099, 0.010) |
| Urbanicity level 2 | -0.028 (-0.079, 0.022) | -0.005 (-0.060, 0.051) | -0.056 (-0.128, 0.016) | -0.014 (-0.096, 0.068) | -0.107 (-0.166, -0.049)** |
| Urbanicity level 3 | 0.011 (-0.052, 0.075) | -0.086 (-0.207, 0.034) | 0.052 (-0.017, 0.121) | -0.062 (-0.162, 0.037) | -0.094 (-0.174, -0.015)† |
| Urbanicity level 4 | 0.010 (-0.062, 0.083) | -0.078 (-0.168, 0.012) | -0.040 (-0.100, 0.020) | -0.091 (-0.159, -0.022)* | -0.007 (-0.068, 0.054) |
| Urbanicity level 5 | 0.011 (-0.022, 0.043) | -0.022 (-0.066, 0.022) | -0.029 (-0.080, 0.021) | -0.006 (-0.066, 0.054) | -0.079 (-0.134, -0.024)* |

Note: † indicates $p < 0.05$; * indicates $p < 0.01$; ** indicates $p < 0.001$; *** indicates $p < 0.0001$; each cell indicates $\beta$ value, 95% CI, $p$ value.

Table S14. **Regression results of association between the local parks and infection rate across five time periods and five urbanicity levels.**

| **Local parks** | **Period 1** | **Period 2** | **Period 3** | **Period 4** | **Period 5** |
|---|---|---|---|---|---|
| Urbanicity level 1 | -0.023 (-0.127, 0.082) | 0.062 (-0.008, 0.131) | -0.021 (-0.051, 0.008) | 0.007 (-0.019, 0.033) | -0.011 (-0.043, 0.020) |
| Urbanicity level 2 | -0.025 (-0.122, 0.073) | 0.071 (-0.035, 0.176) | -0.145 (-0.282, -0.007)† | 0.026 (-0.132, 0.184) | -0.048 (-0.161, 0.065) |
| Urbanicity level 3 | -0.009 (-0.129, 0.110) | -0.002 (-0.236, 0.231) | -0.110 (-0.238, 0.019) | 0.181 (-0.017, 0.380) | -0.006 (-0.166, 0.155) |
| Urbanicity level 4 | 0.018 (-0.203, 0.238) | -0.084 (-0.358, 0.190) | 0.029 (-0.161, 0.219) | 0.163 (-0.054, 0.379) | 0.069 (-0.126, 0.265) |
| Urbanicity level 5 | -0.001 (-0.069, 0.067) | -0.012 (-0.103, 0.080) | -0.082 (-0.187, 0.022) | -0.117 (-0.243, 0.008) | -0.090 (-0.206, 0.026) |

Note: † indicates $p < 0.05$; * indicates $p < 0.01$; ** indicates $p < 0.001$; *** indicates $p < 0.0001$; each cell indicates $\beta$ value, 95% CI, $p$ value.

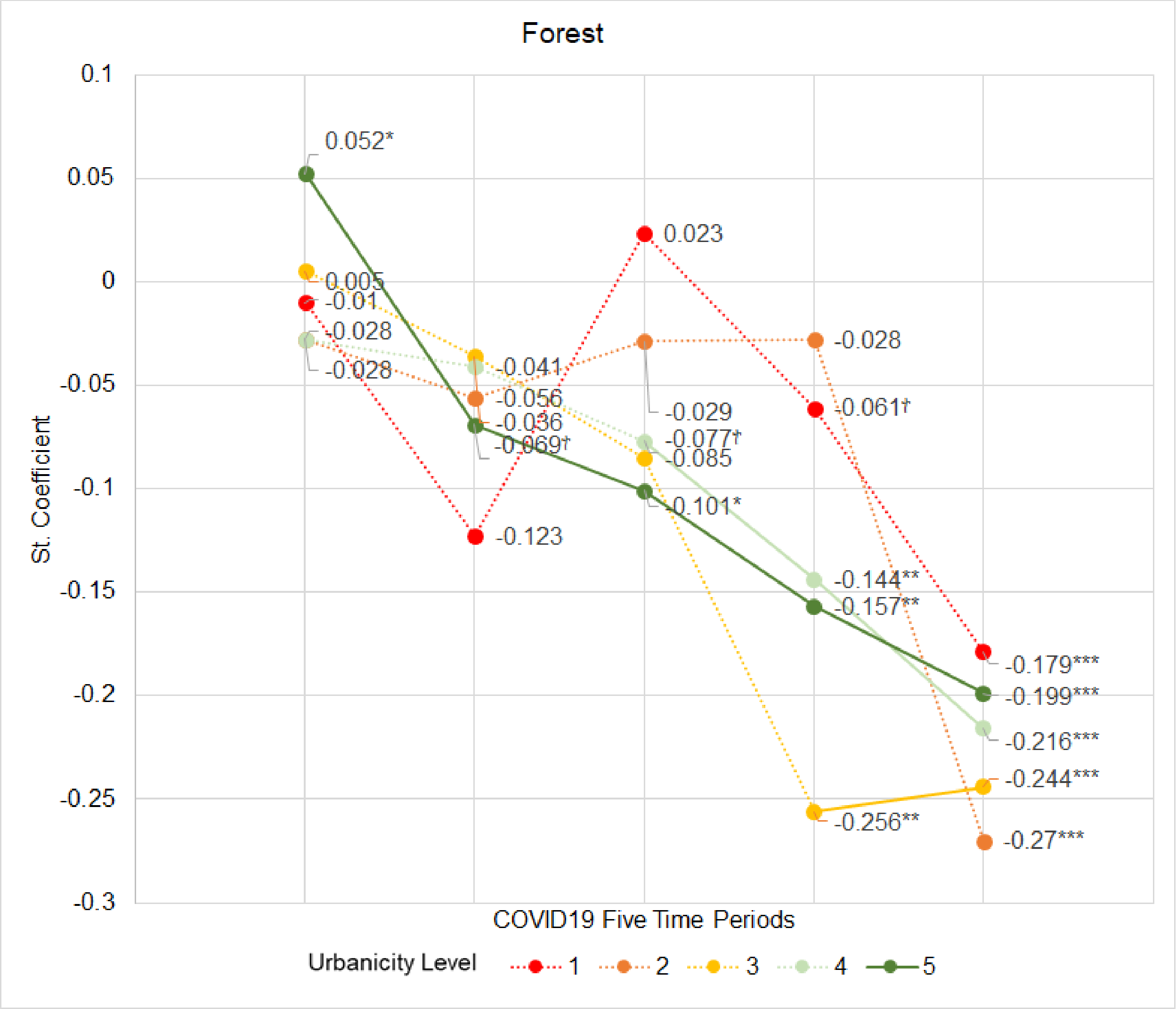


Fig. S1. **Effect of forest on the SARS-CoV-2 infection rate across five time periods and five urbanicity levels.** The number for each point is a $\beta$ value. † indicates $p< 0.05$; * indicates $p < 0.01$; ** indicates $p < 0.001$; *** indicates $p < 0.0001$.

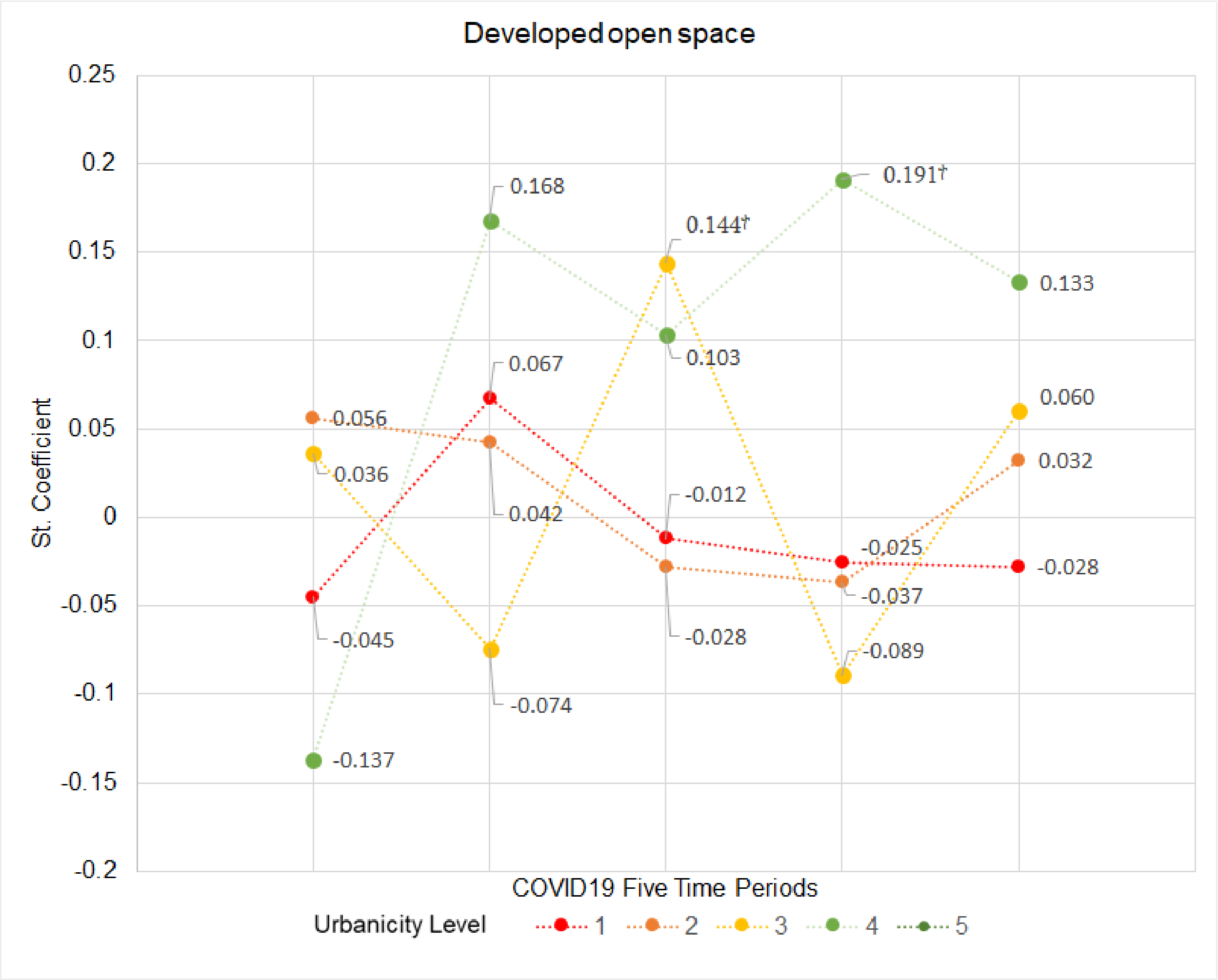


Fig. S2. **Effect of developed open space on the SARS-CoV-2 infection rate across five time periods and five urbanicity levels.** The number for each point is a $\beta$ value. † indicates $p< 0.05$; * indicates $p < 0.01$; ** indicates $p < 0.001$; *** indicates $p < 0.0001$.

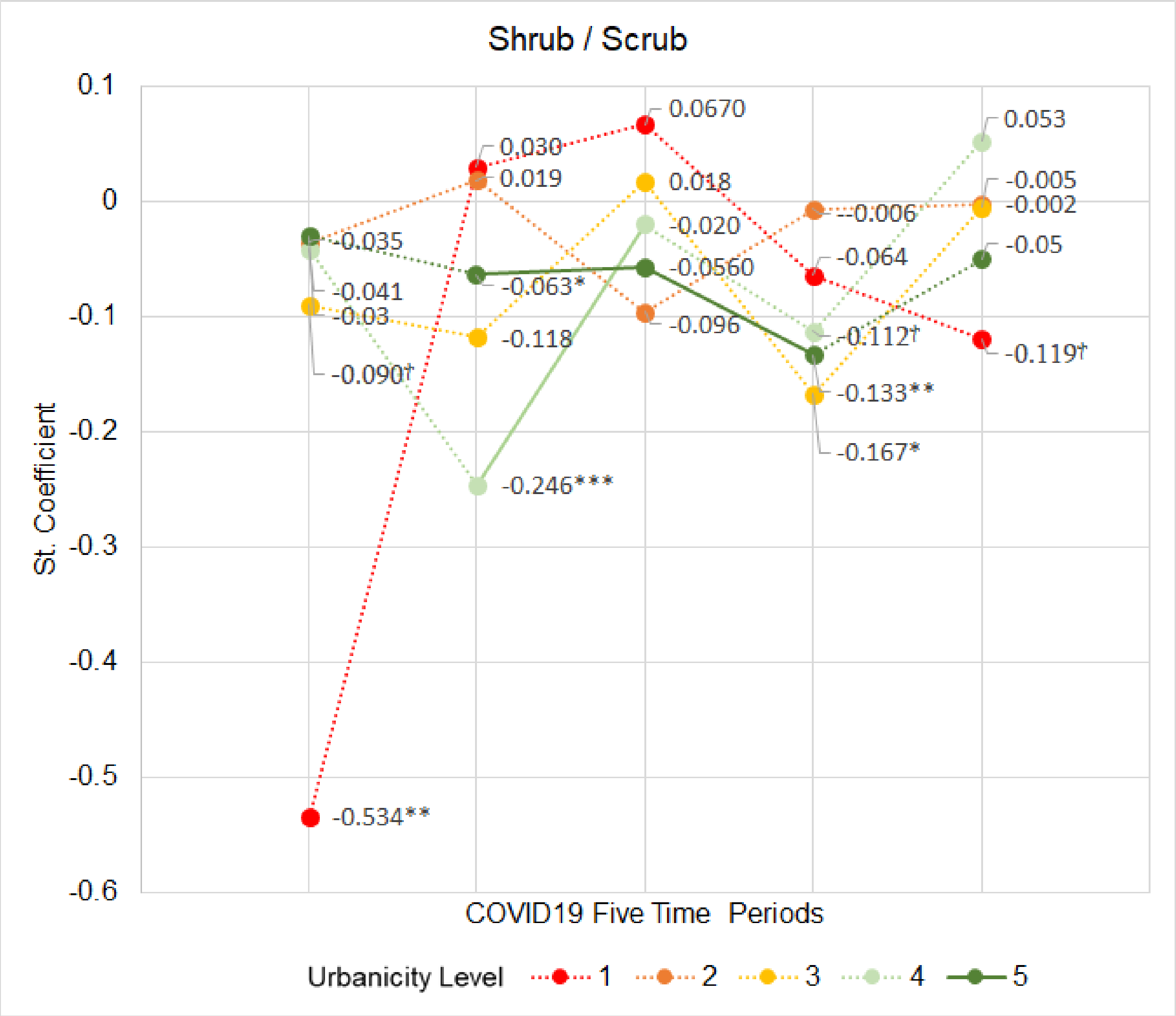


Fig. S3. **Effect of shrub / scrub on the SARS-CoV-2 infection rate across five time periods and five urbanicity levels.** The number for each point is a $\beta$ value. † indicates $p< 0.05$; * indicates $p < 0.01$; ** indicates $p < 0.001$; *** indicates $p < 0.0001$.

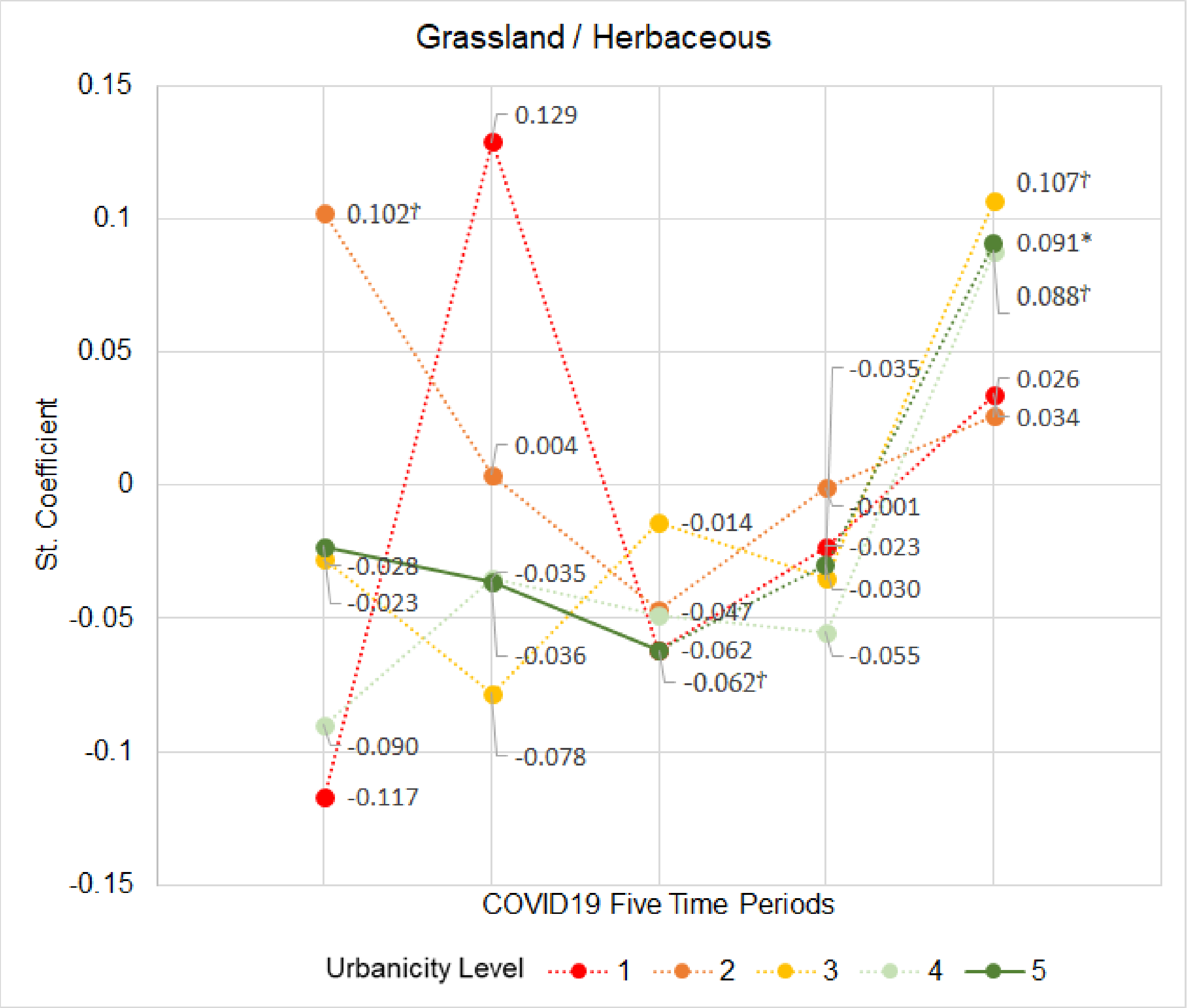


Fig. S4. **Effect of grassland/herbaceous on the SARS-CoV-2 infection rate across five time periods and five urbanicity levels.** The number for each point is a $\beta$ value. † indicates $p$< 0.05; * indicates $p$ < 0.01; ** indicates $p$ < 0.001; *** indicates $p$ < 0.0001.

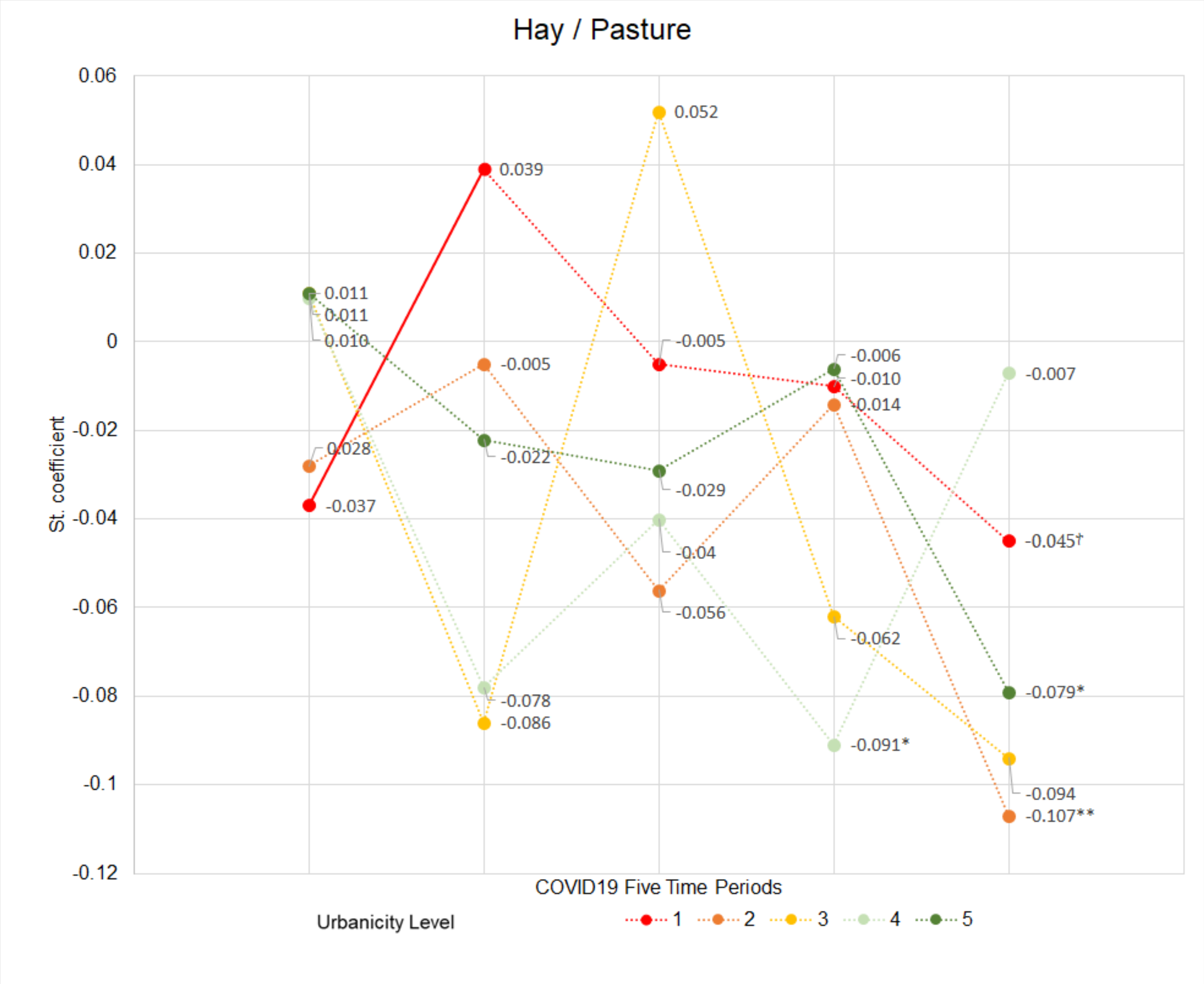


Fig. S5. **Effect of hay / pasture on the SARS-CoV-2 infection rate across five time periods and five urbanicity levels.** The number for each point is a $\beta$ value. † indicates $p< 0.05$; * indicates $p < 0.01$; ** indicates $p < 0.001$; *** indicates $p < 0.0001$.

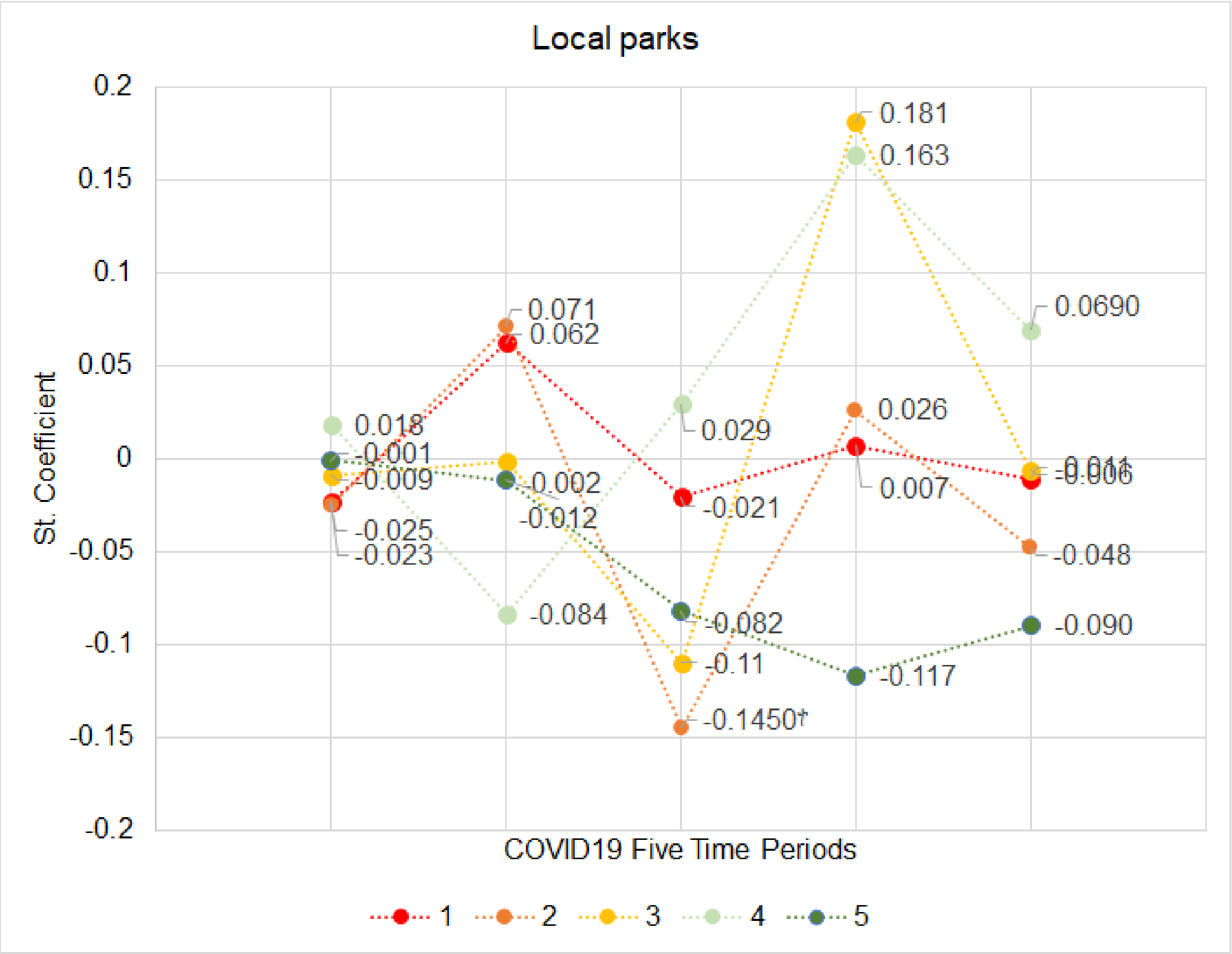


Fig. S6. **Effect of local parks on the SARS-CoV-2 infection rate across five time periods and five urbanicity levels.** The number for each point is a $\beta$ value. † indicates $p < 0.05$; * indicates $p < 0.01$; ** indicates $p < 0.001$; *** indicates $p < 0.0001$.
